# Leveraging fMRI non-stationarity for deep learning classifier training and feature detection to improve schizophrenia diagnosis

**DOI:** 10.1101/2025.10.03.25337252

**Authors:** Madison Lewis, Nicholas Theis, Bowei Ouyang, Konasale M. Prasad

**Affiliations:** Department of Bioengineering, Swanson School of Engineering, University of Pittsburgh, Pittsburgh, PA USA; Department of Psychiatry, University of Pittsburgh, Pittsburgh, PA USA; VA Pittsburgh Healthcare System, Pittsburgh, PA, USA

**Keywords:** <u>Key words</u>: Dynamic functional connectivity, Graph convolutional networks, Saliency analysis, Adolescent-onset schizophrenia, fMRI, Class activation mapping

## Abstract

A neurobiologically-based diagnosis with superior reliability in place of clinical interview-based diagnosis is a primary goal in psychiatry. Dynamic functional connectomes (dFCs) identified using change-point detection applied to functional magnetic resonance imaging (fMRI) data was used to train graph convolutional network (GCN) models to classify persons with psychiatric diagnoses from healthy controls. We examined four samples, adolescent-onset schizophrenia (AOS), adult schizophrenia, major depressive disorder, bipolar disorder, each with healthy controls (HC) for resting state fMRI (rs-fMRI) and working memory task for AOS. Classification accuracy was as high as 89.2% (sensitivity=0.90; specificity=0.88) for adult schizophrenia. The GCNs were further examined to understand which nodes and edges contributed highly to the classification using Class Activation Mapping (CAM) and Integrated Gradients (IG), respectively. CAM and IG analysis were convergent between adult schizophrenia and AOS which included default mode network regions, cerebellum, and sensory regions for rs-fMRI. For working memory, Brodmann area 10 and dorsolateral prefrontal cortex contributed the most towards AOS classification. Applied in a clinical context, post-test probability of accurate classification was 93% for adult-onset schizophrenia using rs-fMRI with a positive test suggesting clinical usefulness of our model. Our results suggest that a combination of deep-learning models and explanatory algorithms can markedly improve diagnostic reliability, offer approaches to objective diagnostic approach, and provide a neurobiological basis for the diagnosis by identifying regions and edges in the networks.

## Introduction

Schizophrenia, with a lifetime risk of approximately 1%^2^ is a chronic brain disorder with profound societal impact, including a 10% risk for completed suicide^3,4^, decreased life span by up to 25 years^5^, and nationwide treatment costs exceeding $340 billion in the US in 2019^6^. Despite intense research, treatment outcomes have not significantly improved over the past 100 years^7^. Since the early classification of mental illnesses into dementia praecox (later called schizophrenia) and manic-depressive illness (currently called bipolar disorder, and others, clinical interviewing and observations have remained the mainstay for the diagnosis. Further, not having a biological basis for these diagnoses has been cited as a reason for poor progress in finding novel and more effective treatments. Since the 1990s^8^, researchers are increasingly relying on magnetic resonance imaging (MRI) to identify brain abnormalities associated with schizophrenia, including volumetric changes in various brain regions^9^, reduced hippocampal volume and shape^10^, enlarged ventricles, and aberrant functional organization^11^, but MRI-based diagnoses is still not realized mainly because pathognomonic radiological markers have not been found. Prior research suggests high-level information processing depends on how brain networks are organized and how they coordinate with each other^12^. Functional connectivity (FC) analysis using machine learning (ML) approaches, given their ability to detect patterns, have been extensively used to uncover patterns of network alterations to better distinguish patient networks from controls and advance the understanding of schizophrenia network pathophysiology^13^.

Deep learning has revolutionized strategies to address the above issues in psychiatry^14^. Among deep learning approaches, convolutional neural networks (CNN) and their spatially generalized counterpart, graph convolutional networks (GCN), have shown particular promise. GCNs accept high dimensional connectivity matrices as inputs unlike support vector machines (SVM); GCN may increase both the model interpretability and the clinical applicability since network dysconnectivity is associated with schizophrenia. Recent research has demonstrated GCNs’ superiority over SVMs in schizophrenia classification, with one study achieving 85% accuracy in binary classification of schizophrenia and controls using static functional connectivity derived from resting state fMRI (rs-fMRI)^15^. Additionally, GCNs enable feature attribution, facilitating biological interpretation and localization of pathology for potential clinical translation^16,17^. Moreover, low test-retest reliability of psychiatric disorders is well known^18,19,20^. Inter-rater reliability for psychotic disorders among clinicians can be as low as 66%^21^ and κ=0.22 for schizoaffective disorder^22^, making classification via network features attractive and meaningful. In our previous study using UK Biobank data, the GCN model more successfully predicted psychosis-spectrum disorders in the community^16^ than unstructured interviews^23^, but we were unable to classify the subset of schizophrenia sample due to small sample size.

In this study, we examined adult schizophrenia, from the same UK Biobank sample, as well as adolescent-onset schizophrenia (AOS) because AOS is understudied and a more severe form of schizophrenia^24^ with poorer long-term outcomes^24^ and more severe cognitive deficits^24,25^ especially in working memory^26,27^ and executive function^28^ compared to adult-onset schizophrenia. AOS constitutes 12.3% of all schizophrenia^29^ and is phenomenologically continuous with adult-onset schizophrenia^26^. Further, schizophrenia is proposed to be a disorder of neurodevelopment with excessive synaptic pruning contributing to the adolescent onset^30^. We also examined major depressive disorder and bipolar disorder sample to obtain relative estimates of accuracy of classification.

We hypothesized that successful GCN training and prediction of case control status is possible using dynamic functional connectomes (dFCs) instead of static FC as we did previously^16^. Change-point detection method was applied to the fMRI time-series for each subject to detect multiple, but unique, FCs for each subject. Recent literature on time varying brain functional connectivity posits that four “c-modes,” or coactivation, modes make up the human brain’s resting state fMRI activity^63^ and supports the assumptions of dynamic fMRI analysis. Cross-covariance isolate detect (CCID) uses the isolate-detect principle^31^ to estimate the number and temporal positions of change-points^32^, creating data-driven and more biologically relevant dFCs than methods such as sliding window. For the working memory task data, we hypothesized a greater proportion of change-points would be in the predicted windows among HC, and conversely a higher proportion of change-points in the unpredicted windows among AOS. Previous studies have trained models on larger static FC datasets of adult schizophrenia^15,33^, and our models were hypothesized to meet or exceed these accuracies. We also expect the regional and edge-wise saliency results to be similar between our AOS and adult schizophrenia samples, thereby validating both models, the robustness of the GCN training, and providing a basis for biological interpretation.

To test these hypotheses, we examined the use of GCNs in a prediction-accuracy approach for classifying dFCs derived from rs-fMRI in AOS, adult schizophrenia, and a transdiagnostic sample consisting of schizophrenia, bipolar disorder, and major depressive disorder, and localizing schizophrenia-associated changes in the brain that contributed to the classification. Since schizophrenia is associated with deficits in working memory^34^ we included an N-back task-positive fMRI sample. We begin with converting the preprocessed fMRI data into regionally averaged timeseries, using the multimodal Human Connectome Project (HCP) Glasser atlas^35^ to define the cortical parcellation, in conjunction with the standard FreeSurfer subcortical structures^36^. We then identified dFCs using change-point detection^31^, which results in multiple functional networks for each subject boosting the network sample size and making it sufficient for GCN model training^37^. After training the GCN models, the model accuracies across different samples were compared and salient feature detection algorithms – class activation mapping (CAM) and integrated gradients (IG) – were deployed. These salient features were compared within a given model (for example the similarity of AOS-CAM to AOS-IG). The salient features were also compared between patient and control groups, as well as between adult schizophrenia and AOS to assess the similarity between them.

## Results

### Sample Characteristics

Four samples were used in this analysis: three samples were from the UK Biobank, adult schizophrenia, bipolar disorder, and major depressive disorder, the latter three groups were combined to form a transdiagnostic group, and one from a locally acquired sample of AOS subjects. The UK Biobank samples consisted of resting-state only whereas AOS sample had resting-state and N-back working memory task fMRI data. Altogether we tested our hypotheses on 6 different datasets.

Age and sex were not significantly different between patient and control groups in any of the samples except for nominally significant sex distribution in the UKB-Bipolar group (**Table 1**). The dFCs were generated using the time series between two adjacent change-points identified by the change-point detection method (**Figure 1**). Each subject had multiple change-points (**Table 1**).

**Table 1:**
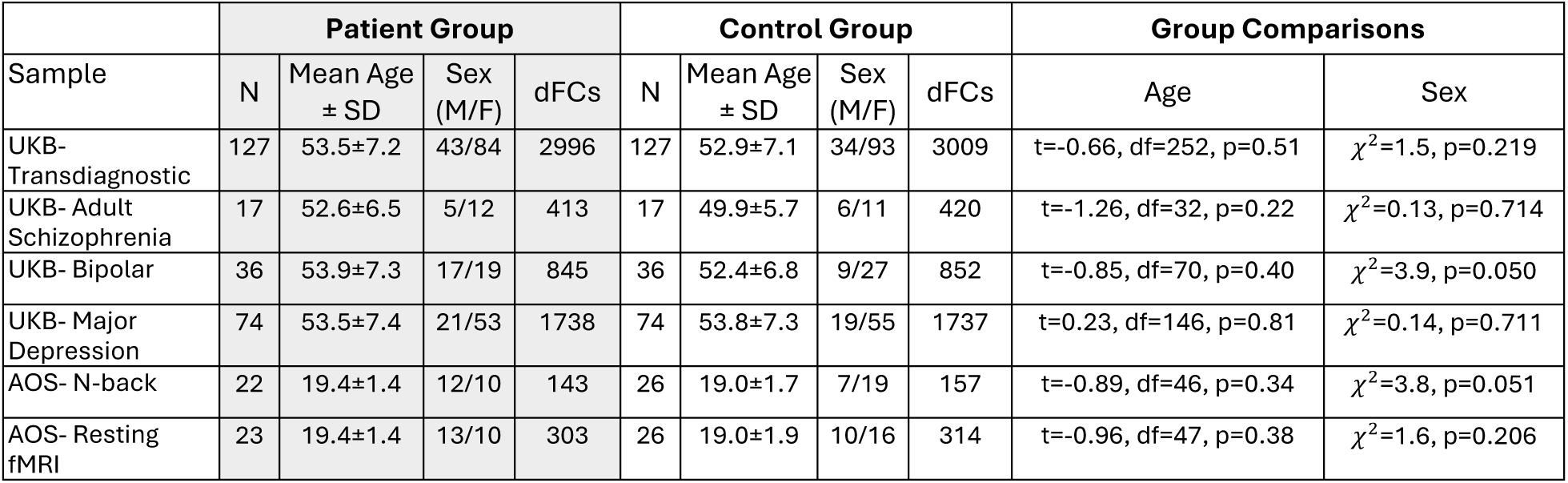
Demographic characteristics for each sample used in the study. Legend: SD Standard deviation; dFCs Number of dynamic functional connectomes.

| Sample | Patient Group |  |  |  | Control Group |  |  |  | Group Comparisons |  |
| --- | --- | --- | --- | --- | --- | --- | --- | --- | --- | --- |
|  | N | Mean Age<br>± SD | Sex<br>(M/F) | dFCs | N | Mean Age<br>± SD | Sex<br>(M/F) | dFCs | Age | Sex |
| UKB- Transdiagnostic | 127 | 53.5±7.2 | 43/84 | 2996 | 127 | 52.9±7.1 | 34/93 | 3009 | t=-0.66, df=252, p=0.51 | $\chi^2=1.5$, p=0.219 |
| UKB- Adult Schizophrenia | 17 | 52.6±6.5 | 5/12 | 413 | 17 | 49.9±5.7 | 6/11 | 420 | t=-1.26, df=32, p=0.22 | $\chi^2=0.13$, p=0.714 |
| UKB- Bipolar | 36 | 53.9±7.3 | 17/19 | 845 | 36 | 52.4±6.8 | 9/27 | 852 | t=-0.85, df=70, p=0.40 | $\chi^2=3.9$, p=0.050 |
| UKB- Major Depression | 74 | 53.5±7.4 | 21/53 | 1738 | 74 | 53.8±7.3 | 19/55 | 1737 | t=0.23, df=146, p=0.81 | $\chi^2=0.14$, p=0.711 |
| AOS- N-back | 22 | 19.4±1.4 | 12/10 | 143 | 26 | 19.0±1.7 | 7/19 | 157 | t=-0.89, df=46, p=0.34 | $\chi^2=3.8$, p=0.051 |
| AOS- Resting fMRI | 23 | 19.4±1.4 | 13/10 | 303 | 26 | 19.0±1.9 | 10/16 | 314 | t=-0.96, df=47, p=0.38 | $\chi^2=1.6$, p=0.206 |

**Figure 1:**
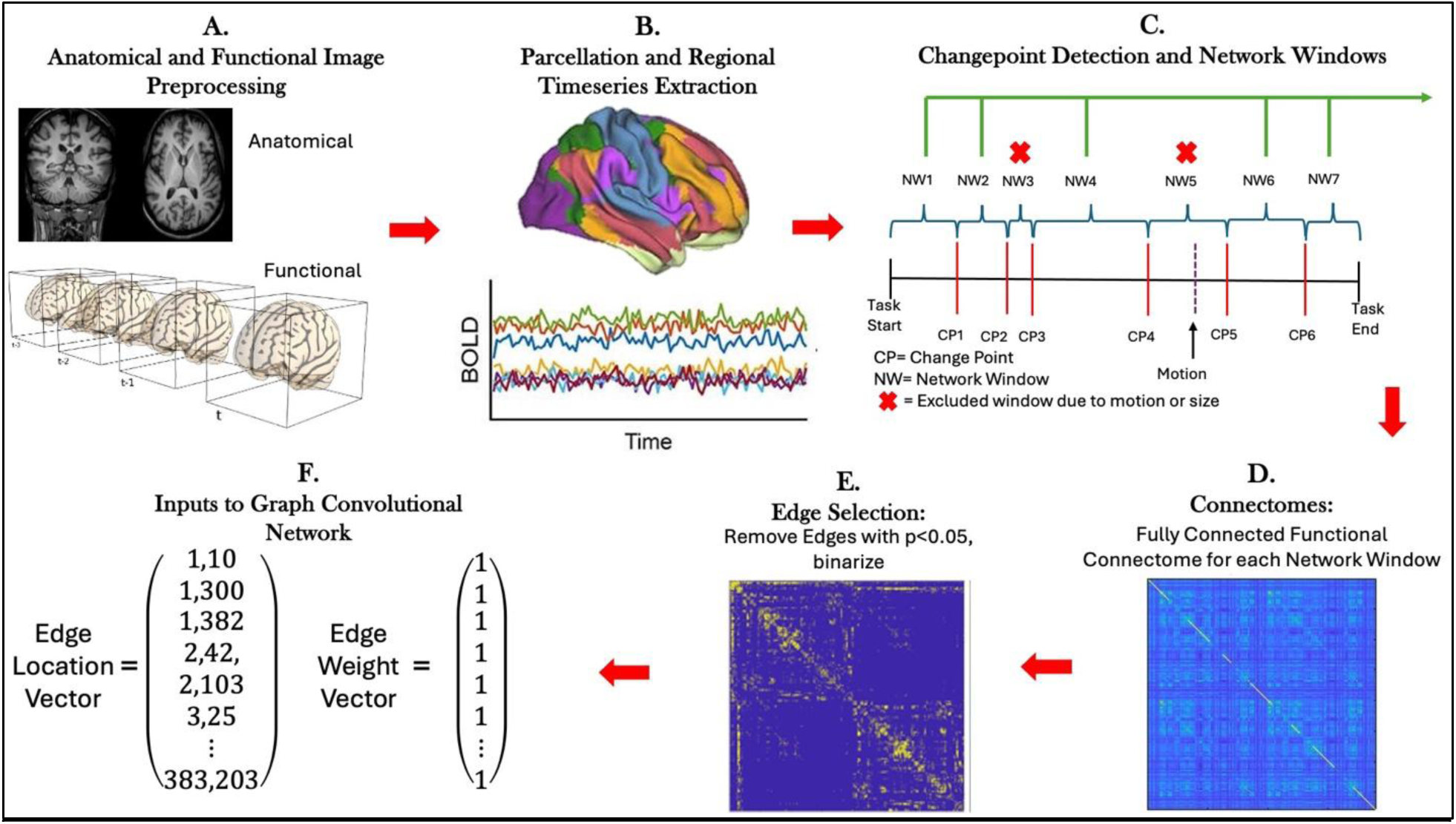
Generation of GCN Inputs. A. Raw anatomical and functional images are preprocessed. B. Anatomical images are parcellated according to the Glasser MMP1 atlas, individual parcellation was registered to the fMRI space, and regional timeseries are extracted for all 383 (subcortical and cortical) regions. C. Change-point (CP) detection using CCID indicated using a red bar. Network windows (NW) are between CPs. Excluded windows are due to window size <10 timepoints (NW3) or suprathreshold motion (NW5) (purple dashed bar). D. Dynamic functional connectome built using pairwise correlations of BOLD signal between two regions for the window between two CCID CPs. E. Edge selection method to create unweighted sparse graphs. F. For inputs into the GCN, edge locations were extracted from unweighted adjacency matrices, and all edge weights with significant correlations were set to 1.

### Change-points and Dynamic Functional Connectomes

While the distribution of change points and resulting dFC window lengths for each GCN input set (AOS: N-back task and resting fMRI; four UKB groups: resting fMRI) were highly variable among subjects but were not significantly different in terms of the number of change-points or dFC window length between diagnostic groups and controls (**Figure 2**). The median number of change-points were 14 for AOS N-back task, 15 for AOS resting fMRI, and 23 for UKB resting fMRI. The median duration of the task dFC window in AOS and adolescent controls was 33 seconds. The median duration of resting state dFC windows was 15 secs in both AOS and adolescent controls. The median duration of resting state dFC windows in the UKB across all clinical groups and controls was about 14 seconds.

**Figure 2:**
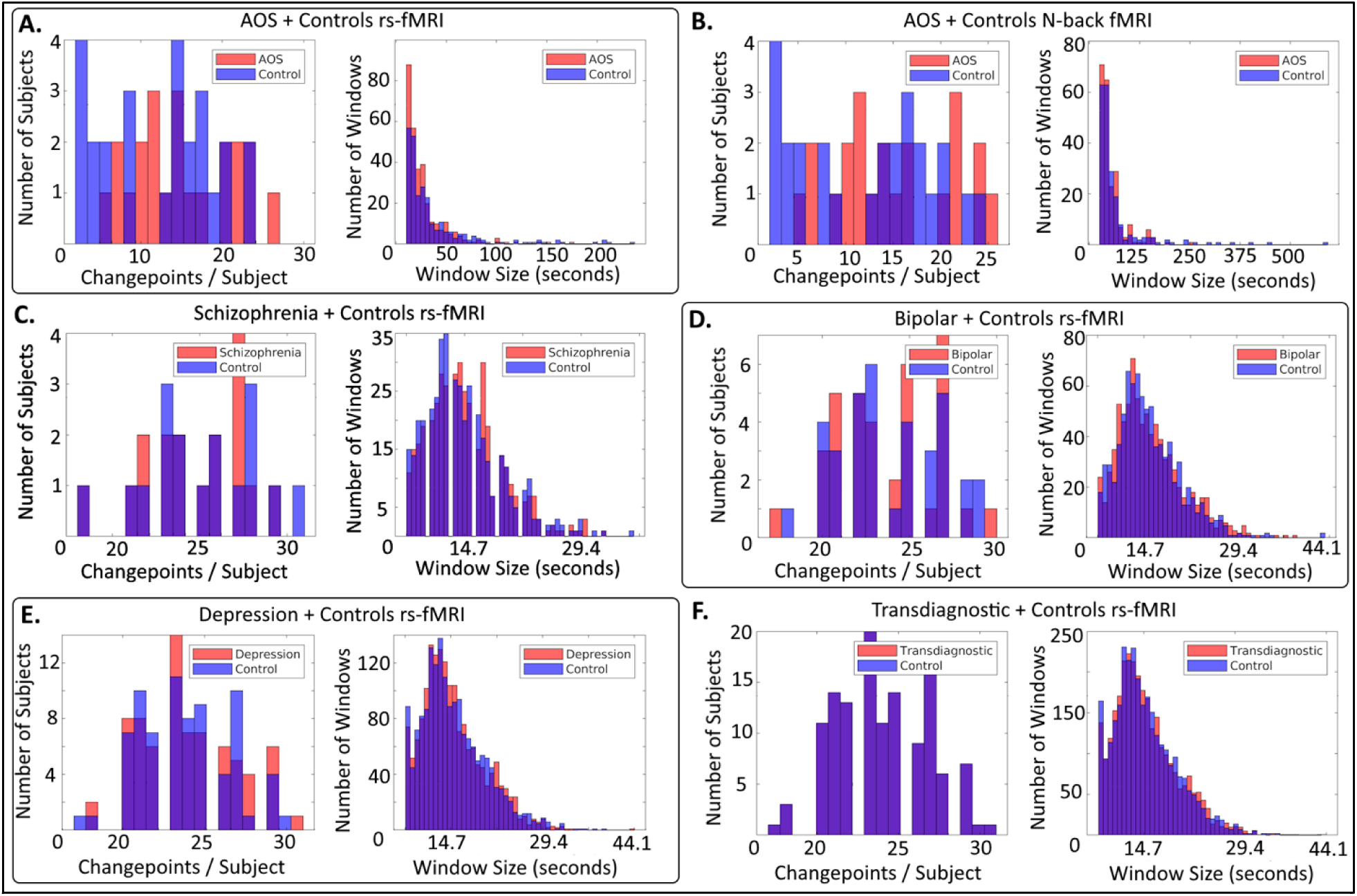
The number of change points detected and the size of dFC windows (in seconds) for each GCN input set. In the task-fMRI (N-back AOS; A) the sampling rate was 2.5 seconds and there were 242 TRs. In the resting state (AOS B) the sampling rate was 1 second and there were 480 TRs. In the UK Biobank datasets (C-F) the sampling rate was 0.735 seconds with 490 TRs.

For the AOS working-memory task data, we compared the proportion of change-points located within 3 TR of either a rest-to-task or task-to-rest transition (predicted window) to the number of change-points not near the rest-to-task or task-to-rest transition (unpredicted window) (Supplemental Fig 1) using χ^2^ test for proportions. For the within-groups analysis, HC had significantly higher proportion of change-points in the predicted windows (4.64%) compared to unpredicted windows (1.62%) for N-back (χ^2^=14.74, p<0.001). A similar pattern was observed for AOS during N-back (predicted window 5.48%, unpredicted window 2.78%, χ^2^=7.3, p<0.001). Between HC-AOS, we noted a significantly higher proportion of TRs in unpredicted windows for N-back among AOS compared to HC (HC 1.62%, EOS 2.78%, χ^2^=16.60, p<0.001).

### Graph Convolutional Network Training

Next, we examined whether the time-series data from the dFCs could be used as inputs to the GCN models to predict the diagnosis. The GCNs were trained for each sample using 5-fold cross validation, except for the transdiagnostic sample which used 10-fold cross validation due to the larger number of networks (**Figure 3**). The number of networks inputted to the GCN models for each sample is shown in **Table 1**.

**Figure 3:**
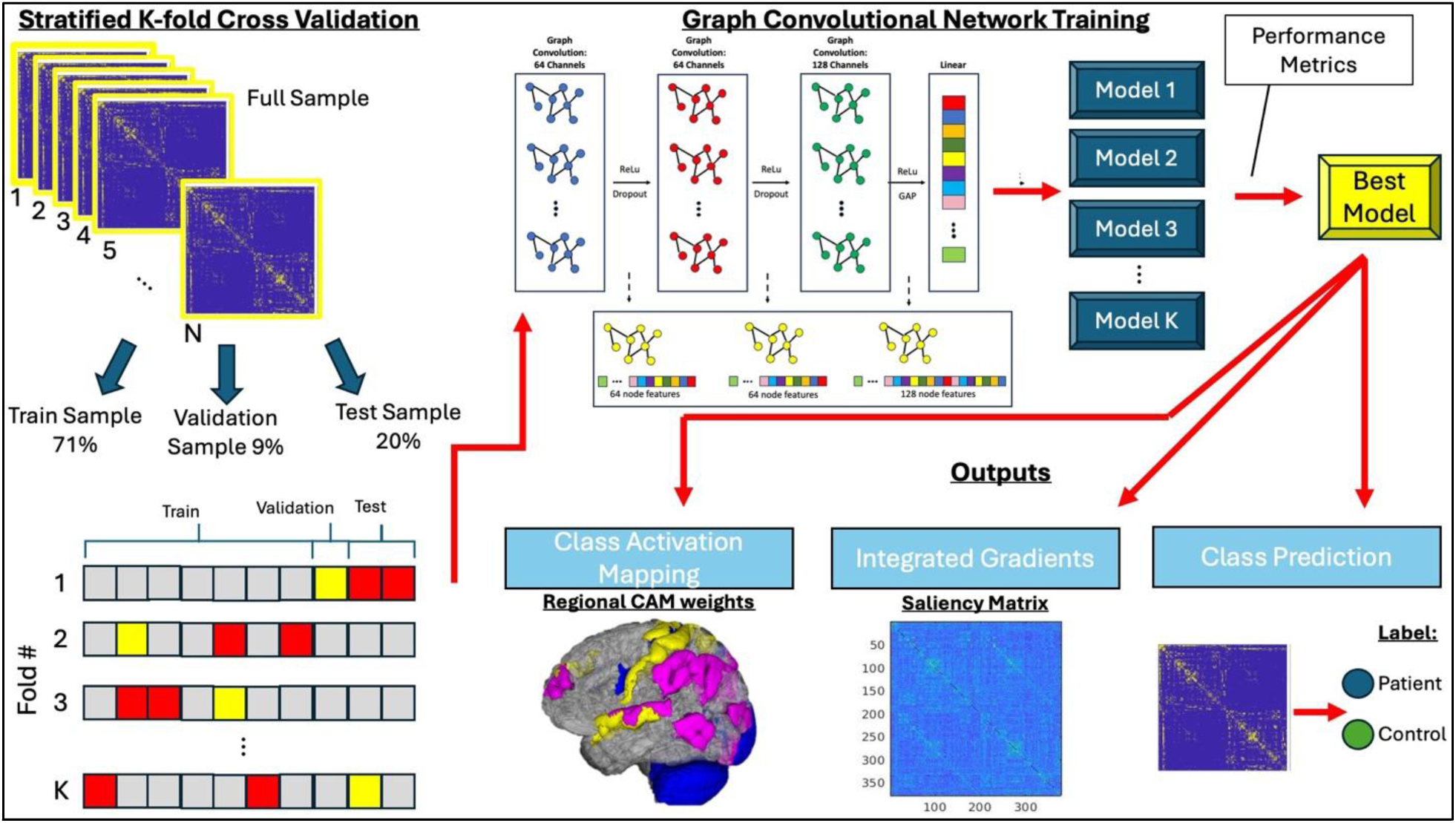
Graph Convolutional Network, Class Activation Mapping, and Integrated Gradients Mixed Model. Using a stratified k-fold cross validation scheme, the data is split into training, validation, and test sets. The training data is then used to train a 3-layer GCN model to classify patients from controls. A model was generated for each fold, and the best model was determined using performance metrics of accuracy, sensitivity, and specificity. The best model was then used for class predication of the entire sample and nodal and edge saliency analysis via class activation mapping and integrated gradients, respectively.

For the UK Biobank rs-fMRI data, the transdiagnostic sample GCN models had 78% average accuracy across 10 folds with 82% best model accuracy (**Table 2**). Of the transdiagnostic subgroups, the adult schizophrenia group performed the best with the best model accuracy of 89% and an average of 87% across 5 folds (**Table 2**) followed by bipolar disorder with an average accuracy of 76% while the best model was 82%. The model performed poorly for major depressive disorder.

**Table 2:** Accuracy, sensitivity, and specificity for each sample’s GCN analysis. Best model and the average across folds accuracy, sensitivity, and specificity are reported. The best model was determined by the highest accuracy.

|  | Best Model Metrics |  |  | Average Metrics Across K-fold Cross Validation |  |  |
| --- | --- | --- | --- | --- | --- | --- |
| Sample | Accuracy | Sensitivity | Specificity | Accuracy | Sensitivity | Specificity |
| UKB- Transdiagnostic | 0.82 | 0.79 | 0.85 | 0.78 ± 0.032 | 0.81 ± 0.030 | 0.75 ± 0.076 |
| UKB- Adult Schizophrenia | 0.89 | 0.90 | 0.88 | 0.87 ± 0.020 | 0.85 ± 0.053 | 0.90 ± 0.028 |
| UKB- Bipolar | 0.82 | 0.82 | 0.83 | 0.76 ± 0.049 | 0.73 ± 0.096 | 0.79 ± 0.043 |
| UKB- Major Depression | 0.75 | 0.78 | 0.71 | 0.69 ± 0.034 | 0.68 ± 0.122 | 0.71 ± 0.121 |
| AOS- N-back | 0.77 | 0.79 | 0.75 | 0.74 ± 0.016 | 0.745 ± 0.100 | 0.73 ± 0.110 |
| AOS- resting fMRI | 0.84 | 0.80 | 0.87 | 0.75 ± 0.067 | 0.70 ± 0.066 | 0.80 ± 0.109 |

For the AOS sample, the models trained using resting-state dFC data achieved 75% average accuracy across 5 folds with the best model achieving 84% accuracy, 80% sensitivity, and 87% specificity (**Table 2**). The GCN models consisting of N-back-derived dFCs performed less well with an average accuracy of 74% with the best model accuracy of 77%. These data suggest that our GCN architecture may be best suited for rs-fMRI-derived dFCs of adult schizophrenia and AOS groups.

### Saliency Analysis for rs-fMRI

Using the best AOS and adult schizophrenia rs-fMRI model, CAM weights were calculated for each *node* (brain region), where the nodes with higher CAM weights contribute proportionately more to correct classification of cases from controls. Similarly, a non-symmetric saliency matrix was calculated using integrated gradients (IG) where each *edge* receives an IG value. For example, an IG value for an edge from node *i* to *j* may not be the same as *j* to *i*. The higher IG values represent edges that are more relevant for classification. Thus, our saliency analysis revealed both the nodes and the edges that contributed to the classification so that the network features defining the diagnoses could be identified.

A spatial representation of the regions on a template brain with a CAM weight in the top 10% (38 regions) for patient and control groups in both rs-fMRI AOS and adult schizophrenia models (**Figure 4**) highlights the similarities/differences between CAM regions in the patient and control groups for the AOS and UKB adult schizophrenia models. All Glasser regions and their CAM weight/rank are in the supplemental spreadsheet.

**Figure 4:**
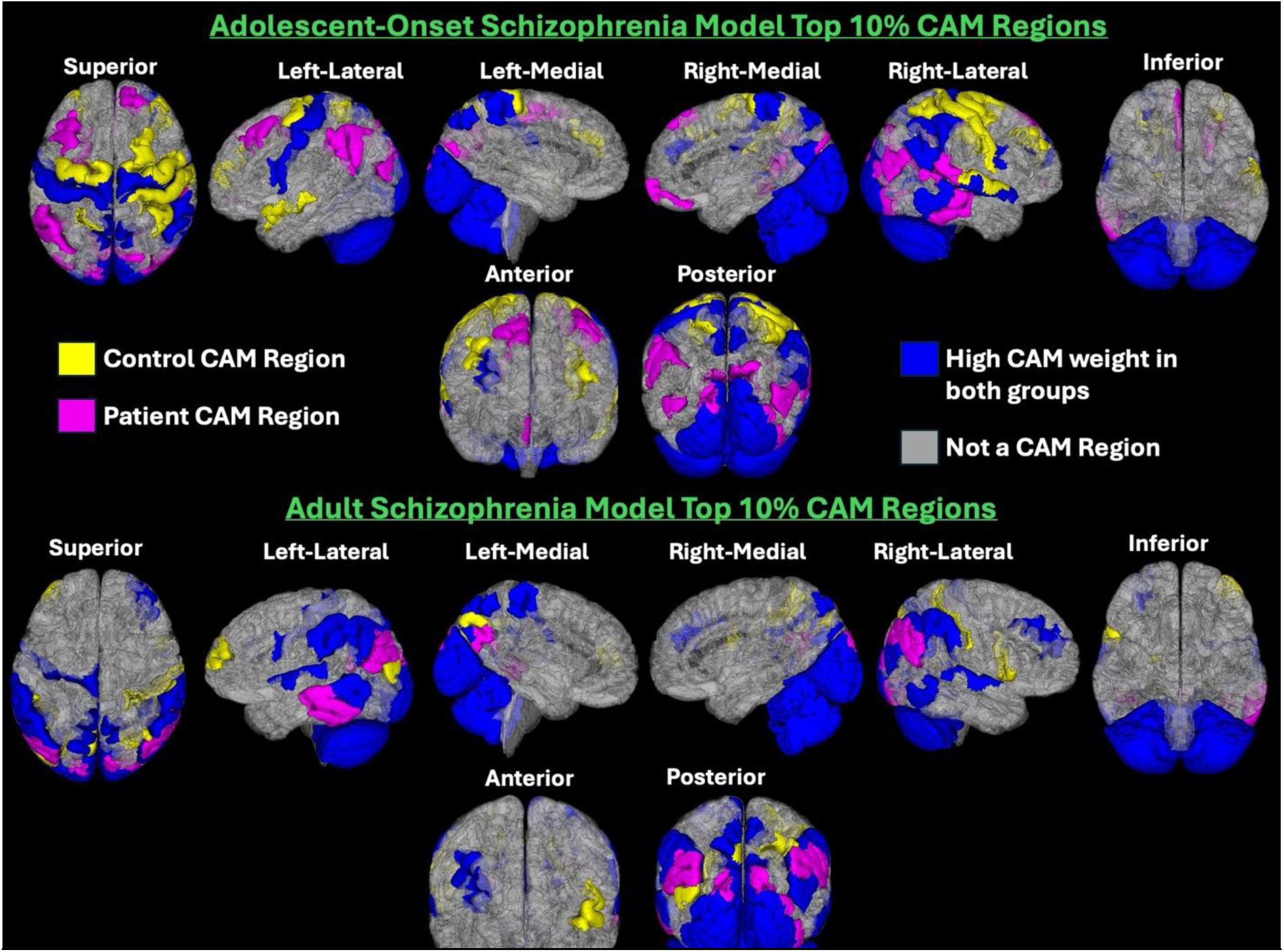
Regions with CAM weights in the top 10*-a comparison between case-control groups. A spatial comparison of the regions with high CAM weights between patient and control groups for the AOS model (top) and adult schizophrenia model (bottom) is shown. Yellow regions indicate regions with high CAM weight which were unique to the control group. Magenta regions were unique to the patient groups. Blue regions have high CAM weights in both the groups. Gray regions were not in the top 10* of CAM weights for either case/control group.

For the GCN model for AOS, of the top 10% of CAM weighted regions, 22 regions (about 58%) were common between AOS and control groups (**Supplemental Table 1**). These were in the subcortical, primary visual, early visual, and somatosensory and motor cortexes. Sixteen regions (42%) were unique to each group and were in the somatosensory and motor, premotor, and superior parietal cortexes for the controls and the primary visual, early visual, and inferior parietal cortexes for the AOS group.

In the adult schizophrenia model, 9 (24%) regions of the top 10% were unique to the class (**Supplemental Table 2**) which is fewer than in the AOS-control comparison. Unique regions to the control group were in dorsolateral prefrontal, somatosensory and motor, and premotor cortexes whereas highly class-activating regions in adult schizophrenia were found in the posterior cingulate cortex, lateral temporal, and early visual cortices. Common regions were located in similar regions as the AOS model (somatosensory, motor, subcortical, the primary visual, early visual, inferior parietal cortexes) and extended beyond them to the posterior cingulate, superior parietal, lateral temporal, auditory association, and dorsolateral prefrontal cortexes (DLFPC).

Although some Glasser sections were both common and unique in these groups, e.g. DLPFC, the HCP parcels within the Glasser sections were different. The HCP parcels unique to AOS within the DLPFC section were areas 8B lateral and 8Av whereas the parcel unique to HC was area 46. HCP parcel common to both AOS and HC within the DLPFC section was area 9-46d. Supplemental table 2 provides information on all common/unique Glasser sections and HCP parcels.

Next, we compared the similarity between CAM regions by comparing the AOS with adult schizophrenia and the young adult controls from the AOS sample with older adult controls from adult schizophrenia sample. The patient groups in the AOS and adult schizophrenia model shared high CAM weighted regions in the left and right cerebellum, right primary visual cortex, left second visual area, and left primary visual cortex (**Figure 5C**). The control groups shared high CAM weighted regions in the left and right cerebellum cortex, left primary visual cortex, right second visual area, right area 2, and right medial area 7A (**Figure 6C**).

**Figure 5:**
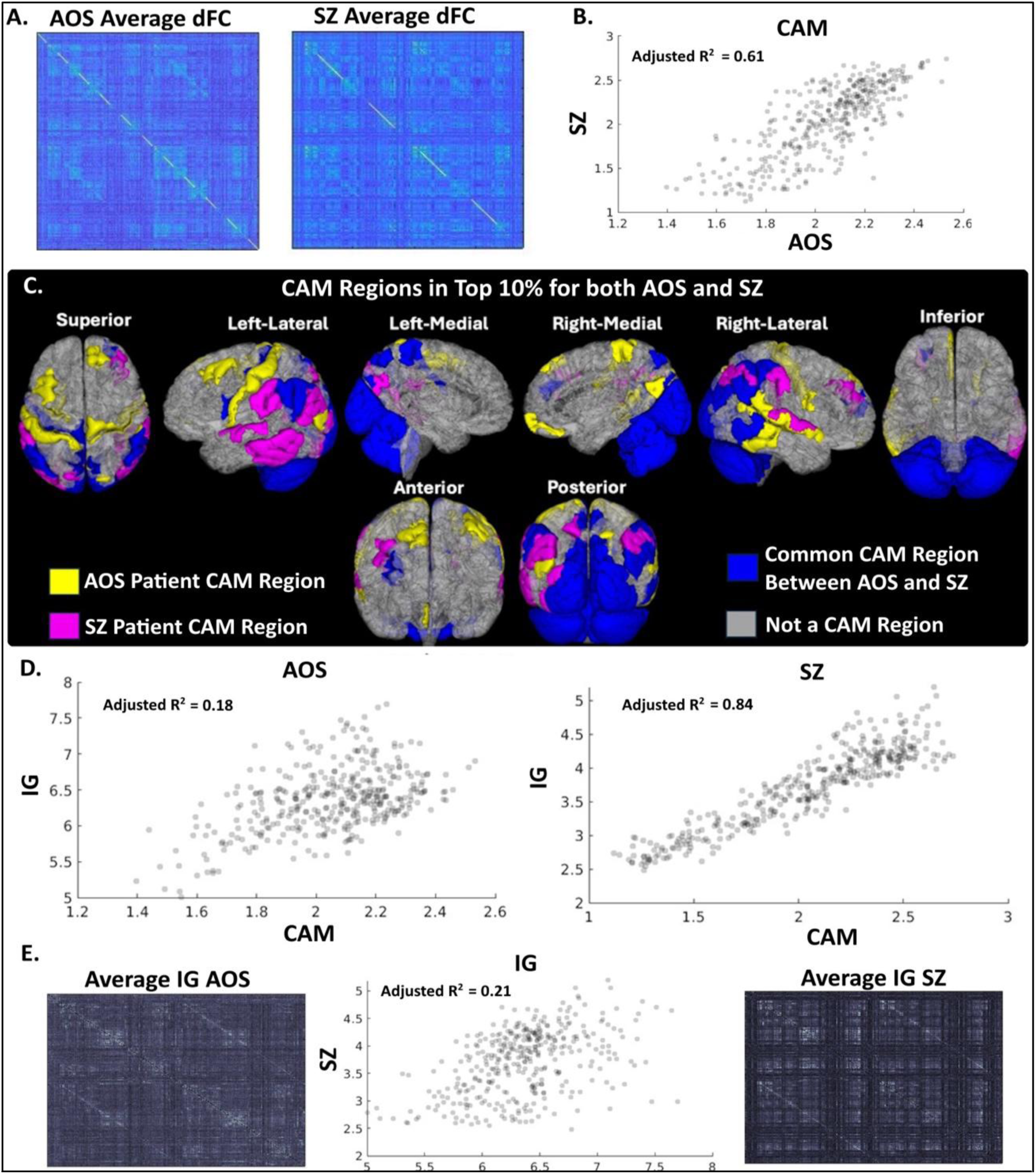
Saliency Analysis of AOS and adult schizophrenia. A. The average weighted dFC network over all dFCs in the AOS rs-fMRI and adult schizophrenia rs-fMRI. B. Scatter plot showing the relationship between CAM weights among all nodes in the AOS and adult schizophrenia classes. C. The top 10% of salient CAM regions plotted on a representative 3D brain with regions in the top 10% for AOS only (yellow), adult schizophrenia only (magenta) and overlapping regions among the top 10% in both groups (blue) for 8 views of the brain. D. Scatter plots showing the relationship between total nodal IG and CAM for each AOS and for adult schizophrenia. E. Visual representation of IG patterns in the networks for AOS (left) and adult schizophrenia (right) as well as a scatter plot total nodal IG between AOS and adult schizophrenia.

**Figure 6:**
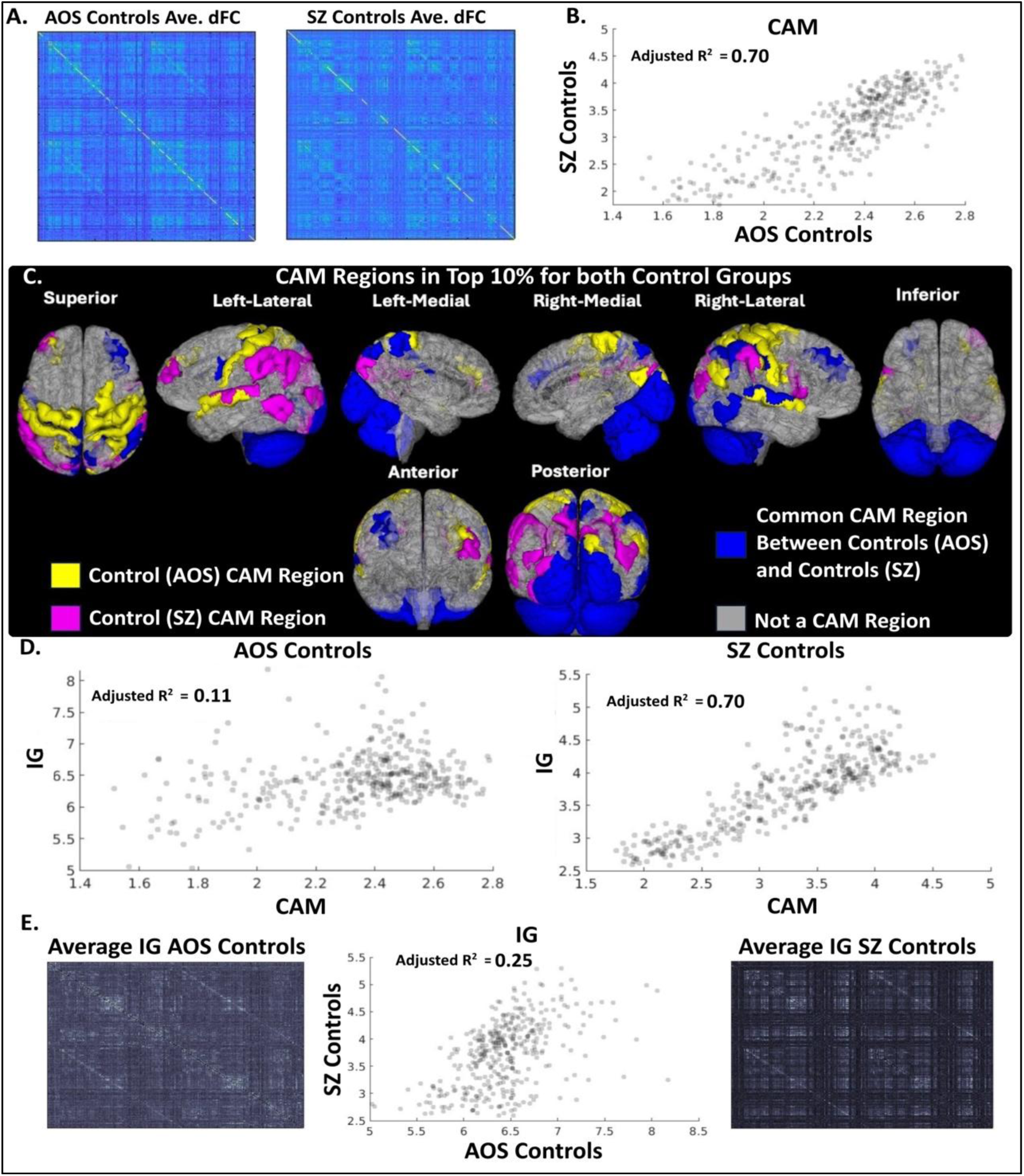
Saliency Analysis of AOS-controls and adult schizophrenia -controls. A. The average weighted dFC network over all dFCs in the AOS-control rs-fMRI and adult schizophrenia -control rs-fMRI. B. Scatter plot showing the relationship between CAM weights among all nodes in the AOS-control and adult schizophrenia-control classes. C. The top 10% of salient CAM regions plotted on a representative 3D brain with regions in the top 10% for AOS-control only (yellow), adult schizophrenia -control only (magenta) and overlapping regions among the top 10% in both groups (blue) for 8 views of the brain. D. Scatter plots showing the relationship between total nodal IG and CAM for each AOS-controls and for adult schizophrenia-controls. E. Visual representation of IG patterns in the networks for AOS-controls (left) and adult schizophrenia-controls (right) as well as a scatter plot total nodal IG between AOS-controls and adult schizophrenia-controls.

After qualitative comparison of the top 10% of CAM regions, we quantitatively compared all the CAM weights for the concordance of CAM weights between the patient groups and between control groups separately in the two independently trained models. The correlation of average CAM weights between the two patient groups (r=0.78, p=1.39e-79) and between two control groups (r=0.84, p=9.65e-100) were significant with a high agreement for the CAM weights for patients (R^2^=0.61, **Figure 5B**) and controls (R^2^=0.70, **Figure 6B**). These highly correlated and statistically similar weights suggest that the same regions influence the classification of patients and controls in two separately trained GCN models, although the rank order of influence varied between samples. This demonstrates the robustness of GCN models and CAM’s ability to detect salient regions and edges contributing to classification that may also be associated with schizophrenia pathophysiology.

Additionally, we investigated whether there was a relationship between the CAM weights and the regional sum of edge saliency values, i.e. sum of all edge saliency values for each node. For all patient and all control groups, we correlated the CAM weights with the regional sum of the edge saliency values and calculated the similarity using adjusted R^2^. For the resting-state UKB adult schizophrenia model, the CAM weights and the regional sum of edge saliency values were highly correlated for both patients (r=0.92, p=1.14e-151) and controls (r=0.84, p=1.31e-9) with a high agreement for both adult schizophrenia model for patients (adjusted R^2^= 0.85) (**Figure 5D, right**) and controls (adjusted R^2^=0.70) (**Figure 6D**, right). This suggests a high degree of spatial correspondence between CAM and IG. To further demonstrate this, we investigated if regions with CAM weights in the top 5% also had regional sum of edge saliency in the top 5%. For the adult schizophrenia group, six regions were in the top 5% in both CAM weights and regional sum of edge saliency: left Auditory 4 Complex (Auditory Association Cortex), left Area PGs, left Area PGi, right Area PF Complex, left Area PFm Complex, and right Area PGi (Inferior Parietal Cortex) suggesting that the nodes and edges of these nodes drove the classification.

The resting-state AOS model showed significant correlations with medium effect size between the nodal CAM weights and sum of edge IG values for the patient (r=0.49, p=3.510e-10) and control groups (r=0.33, p=6.55e-11). However, we observed a wider spread and smaller adjusted R^2^ values for the AOS model than the UKB adult schizophrenia model with adjusted R^2^= 0.18 (**Figure 5D, left**) and R^2^=0.11 (**Figure 6D**, **left**) for AOS and controls, respectively suggesting that higher number of areas were more variable compared to a small proportion that were similar. This may be due to the adolescent brain maturation that is heterochronous compared to the adult brains. Since the AOS model had a wider spread, we searched for the top 10% of CAM weights and regional sum of edge saliency values. The AOS group had two regions in the top 10%: right Area 10v (Anterior Cingulate and Medial Prefrontal Cortex) and right Area 8B Lateral (DLPFC).

In the same way as the CAM weight, we compared the nodal sum of edge saliency weights between patient and control groups across the models. The sum of the edge saliency values showed a significant correlation with medium effect size like the CAM weights for patient (r=0.46, p=1.83e-21) and control (r=0.50, p=1.54e-25) groups but relatively lower agreement than the CAM weights between patient groups (R^2^=0.21, **Figure 5E**) and controls (R^2^=0.25, **Figure 6E**). These similarities across models further demonstrate the validity of the GCN models and the identification of functional connections which could be associated with schizophrenia.

In a similar way to the direct comparison of the top 10% of CAM regions, we directly compared the top ten salient connections in all groups across both models (**Table 3**). Controls in both AOS and adult schizophrenia models had 4 commissural (between hemisphere) and 6 associational (within hemisphere) connections whereas AOS had 6 commissural and 4 associational edges. Adult schizophrenia model showed 9 associational edges with only 1 commissural edge. The AOS model revealed three identical highly salient connections between AOS and its controls whereas the adult schizophrenia model had four identical connections with its controls (**Table 3**).

**Table 3:** Top 10 salient connections for each patient and control group in both AOS and adult schizophrenia models. Red font indicates the same connection in both patients and controls and is shown at the top of each column. Connection based on Glasser sections of the brain is also shown for a spatial representation of the salient edges. Edge type labelled each connection as either commissural, interhemispheric connection, or associational, a connection within the same hemisphere. <u>Legend</u>: ACMPC-Anterior Cingulate and Medial Prefrontal Cortex, DLPFC-Dorsolateral Prefrontal Cortex, DSVC-Dorsal Stream Visual Cortex, EVC-Early Visual Cortex, IFOC-Insular and Frontal Opercular Cortex, IPC-Inferior Parietal Cortex, OPFC-Orbital and Polar Frontal Cortex, PCC-Posterior Cingulate Cortex, PVC-Primary Visual Cortex, SMC-Somatosensory and Motor Cortex, SUB-subcortical.

| Top 10 Salient Connections of Adolescent-Onset Schizophrenia Classification Model |  |  |  |  |  |
| --- | --- | --- | --- | --- | --- |
| Controls |  |  | Adolescent-Onset Schizophrenia |  |  |
| Connection | Glasser Section | Edge Type | Connection | Glasser Section | Edge Type |
| L Polar 10p→L Area 10v | OPFC→ACMPC | Associational | L Polar 10p→L Area 10v | OPFC→ACMPC | Associational |
| L Area 10v→L Polar 10p | ACMPC→OPFC | Associational | L Area 10v→L Polar 10p | ACMPC→OPFC | Associational |
| L Area 10d→Area 10d | OPFC→OPFC | Commissural | L Area 10d→R Area 10d | OPFC→OPFC | Commissural |
| L Caudate→R Caudate | SUB→SUB | Commissural | L Pallidum→L Putamen | SUB→SUB | Associational |
| R Caudate→L Caudate | SUB→SUB | Commissural | L Second Visual Area→L Primary Visual Cortex | EVC→PVC | Associational |
| L Frontal Opercular Area 2→L Frontal Opercular Area 3 | IFOC→IFOC | Associational | L Orbital Frontal Complex→R Orbital Frontal Complex | OPFC→OPFC | Commissural |
| R Area 9 Middle→R Area 9 anterior | ACMPC→DLPFC | Associational | R Area 10d→L Area 10d | OPFC→OPFC | Commissural |
| L Area 9 Middle→L Area 10d | ACMPC→OPFC | Associational | R Area 10v→L Area 10v | OPFC→ACMPC | Commissural |
| L Polar 10p→R Polar 10p | OPFC→OPFC | Commissural | R Area p32→L Area 10r | ACMPC→ACMPC | Commissural |
| R Area 8B Lateral→R Area 9 anterior | DLPFC→DLPFC | Associational | R Orbital Frontal Complex→L Orbital Frontal Complex | OPFC→OPFC | Commissural |
| Top 10 Salient Connections of Adult Schizophrenia Classification Model |  |  |  |  |  |
| Controls |  |  | Adult Schizophrenia |  |  |
| Connection | Glasser Section | Edge Type | Connection | Glasser Section | Edge Type |
| R Area 9 Middle→L Area 9 Middle | ACMPC→ACMPC | Commissural | R Area 9 Middle→L Area 9 Middle | ACMPC→ACMPC | Commissural |
| L Area 10d→L Area 9 Middle | OPFC→ACMPC | Associational | L Area 10d→L Area 9 Middle | OPFC→ACMPC | Associational |
| R Area PGs→R Area PGi | IPC→IPC | Associational | R Area PGs→R Area PGi | IPC→IPC | Associational |
| L Area 9 Middle→L Area 10d | ACMPC→OPFC | Associational | L Area 9 Middle→L Area 10d | ACMPC→OPFC | Associational |
| R Area 9 Middle→R Area 8B Lateral | ACMPC→DLPFC | Associational | L Area 9 Middle→L Area 9 anterior | ACMPC→DLPFC | Associational |
| R Area 8B Lateral→R Area 9 Middle | DLPFC→ACMPC | Associational | R Area PGi→R Area PGs | IPC→IPC | Associational |
| R Area ventral 23 a+b→L Area ventral 23 a+b | PCC→PCC | Commissural | L Area 9 Middle→L Area PGi | ACMPC→IPC | Associational |
| L Area ventral 23 a+b→R Area ventral 23 a+b | PCC→PCC | Commissural | L Third Visual Area→L Area V3A | EVC→DSVC | Associational |
| L Area IntraParietal 1→R Area IntraParietal 1 | IPC→IPC | Commissural | L Area 2→L Area 1 | SMC→SMC | Associational |
| L Area PGs→L Area 8Ad | IPC→DLPFC | Associational | R Primary Sensory Cortex→R Primary Motor Cortex | SMC→SMC | Associational |

### Working Memory Performance in Schizophrenia and Saliency Analysis

Working memory was assessed using the in-scanner N-back task data. The processing time was significantly prolonged among AOS overall as well as in the 2-back and 1-back but not in the 0-back condition (**Supplemental Table 3B**). However, the accuracy of performance on overall N-back, 0-back, 1-back, or 2-back did not show differences between AOS and adolescent controls (**Supplemental Table 3A**). All specific salient brain regions and connections are shown in **Supplemental Table 4**.

## Discussion

Six patient-control samples of dFCs generated using data-driven change-point detection method were provided as inputs to train separate GCN models to test whether the dFCs can classify patients and controls and test its reproducibility across these samples (**Figures 1 and 3**) and whether the resulting classification had neurobiological basis using the saliency analysis through feature extraction of the neural networks. We show that patient-control classifications can be successfully made using the dFCs as inputs to train the GCN models that is better than the average reliability among clinicians and show the nodes and edges that contribute to the classification highlighting the underlying neurobiological basis of classification^38^. This can be further examined in the future studies for subclassification of schizophrenia based on network features.

The minimum number of dFCs used for training across six GCNs was 300 providing adequate sample power for the models (**Table 1**). These binary networks were determined by the significance of the BOLD signal timeseries correlation between pairs of regions that were selected using the multi-modal human brain cortical atlas^35^ and the standard FreeSurfer subcortical structures^31,36^ resulting in approximately 20 dFCs per individual, with variability across the samples but no significant patient-control differences in window number or duration in any set (**Figure 2**). We observed differences in the timing of change-points during N-back task between AOS and controls. The network architectures changed more frequently among AOS during N-back within a task block, whereas network architectures among HC changed more often when the nature of the task changed. These results suggest that connectivity patterns in AOS show unstable or poorly coordinated pattern of regional activations compared to more stable pattern among HC suggesting potential biological significance of dFCs.

Using the dFC networks to train classifiers resulted in moderate to high accuracy depending on the diagnostic group (**Table 2**), with the poorest performing models being trained on the major depression resting state networks (best accuracy of 75%), marginally better performance in the AOS working memory set (77% best model accuracy), and the best performing model having been trained on the adult schizophrenia resting state dataset (89.2% best accuracy). The average accuracy in this model (87.1%) was higher than the accuracies for average model performance in prior publications (85.8%) using larger samples (N=295 schizophrenia patients and N=452 controls pooled from five research centers) and *static* FC^15^ suggesting that using change-point informed dFC may be biologically more appropriate and may augment the sample power since the number of dFCs per subject is used for GCN training rather than the number of subjects. Our study demonstrates that GCN can classify bipolar disorder, major depression, schizophrenia, and AOS using dFCs as inputs with variable accuracies. Modifying the GCN architecture may further improve the accuracies.

Using our model in a clinic may enhance the probability of diagnosing schizophrenia with greater confidence. For adult-onset schizophrenia, an individual presenting with psychotic symptoms in the absence of significant substance use or underlying medical disorders, with a clinician’s mean diagnostic reliability of 65%, the post-test odds of having schizophrenia would be 13.93 and a post-test probability of 93% with a successful classification by the GCN model (“positive test”). When the test is negative, the post-test odds and probabilities of having schizophrenia would be 0.074 and 7%. However, the results were not as encouraging for AOS within the clinic. Since the diagnostic reliability can be low for AOS, assuming a 50% reliability of clinician’s diagnosis, post-test odds and probabilities was 2.96 and 75% for the resting state data with a positive test, and 0.74 and 7%, respectively with a negative test. For the working memory task fMRI dataset, it was even lower with post-test odds and probabilities of 1.52 and 60%, respectively for positive test and 0.09 and 8%, respectively for a negative test. We did not envision this classifier be used in the community to screen for schizophrenia. With a community lifetime prevalence of 1% for schizophrenia, the post-test odds and probabilities were 0.08 and 7%, respectively. With approximately 39% of male and 23% of female patients develop schizophrenia before 19 years of age^29,39^, assuming an average of 31% of 1% prevalence for schizophrenia for AOS as pre-test odds, the post-test odds and probabilities for AOS was 0.02 and 2% for a positive test and 0.001 and 0%, respectively for a negative test for using N-back; for the resting state fMRI data, the post-test odds and probabilities was 0.03 model is likely to be quite successful for adult schizophrenia with resting state data with encouraging results for AOS, when applied in the clinic after a clinician’s evaluation.

An additional focus of our study was to elucidate whether the deep learning-based classification has neurobiological basis that are reproducible in independent data sources. Applying saliency analysis using nodal CAM and IG for edges, we found distinct and overlapping regions and edges. Because the schizophrenia samples are of primary interest, and because the GCNs trained on the adult schizophrenia and AOS inputs achieved the highest classification accuracies for resting state, saliency analysis was performed for the schizophrenia samples. Within each GCN model, for each diagnostic group (i.e. patients or controls), nodal CAM weights were proportional and spatially similar to the nodal sum of IG weights (**Figure 5**). This was more apparent in the adult schizophrenia sample. Comparing the CAM and IG findings between AOS and adult schizophrenia groups derived from separately trained models, CAM weights identified many common and unique regions within patients and controls, a pattern that held but with a smaller effect in terms of total nodal IG (**Figure 6**). These correlations suggest that not only do the saliency methods concur, but that the two independent GCN networks, trained on entirely non-overlapping data, are able to classify with similar accuracy based on the same features. In addition, spatial similarity between CAM and IG also suggest that the identified nodes and their edges together may be contributing to the classification supporting network abnormalities in adult and adolescent-onset schizophrenias.

The locations in the brain of the top 10% of CAM regions for resting state (**Figure 4**) revealed differences and similarities between locations of CAM regions in both AOS and adult schizophrenia and their respective controls. Many regions were in the same Glasser *<u>sections</u>*, but HCP parcels were distinctly different **(Figure 7)**. The Glasser sections were defined in the original publication as collections of geographically contiguous regions that shared common properties such as architecture, task-fMRI profiles, and/or functional connectivity^35^. For example, in the AOS-control comparison, DLPFC Glasser section was both common between AOS and controls but also unique to AOS. However, the parcel that was common between AOS and control was the right area 9-46d but the unique parcels for AOS were the left area 8Av and 8B lateral suggesting that relatively more extensive network contributes to classifying AOS than controls. These parcels within the DLFPC section have different connectivity patterns and serve different functions, e.g. area 9-46d serves goal directed higher-order cognitive processing and conscious planned behavior whereas area 8Av is involved in spatial memory and processing^1^. Similar patterns were present in the parietal, visual, sensory-motor, and other frontal regions (**Supplemental Table 1&2**). The spatial overlap was also similar between adolescent and adult patients between models and between control groups (**Figure 4**). The cerebellum was highly influential for classification of schizophrenia at rest (**Supplemental Table 2**) and in working memory (**Supplemental Table 4**). There is a plethora of evidence associating cerebellar abnormalities with schizophrenia^40^ including functional dysconnectivity in first-episode psychosis^41^, working memory deficts^42^, reduced volumes^43^, and aberrant connectivity of the cortico-cerebellar-thalamic-cortical circuit resulting in loss of “cerebellar coordination of cognitive activity” leading to symptoms associated with psychosis^40^. Since the Glasser atlas used here includes the entire cerebellum as one node, we could not identify specific subregions of cerebellum associated with schizophrenia.

**Figure 7:**
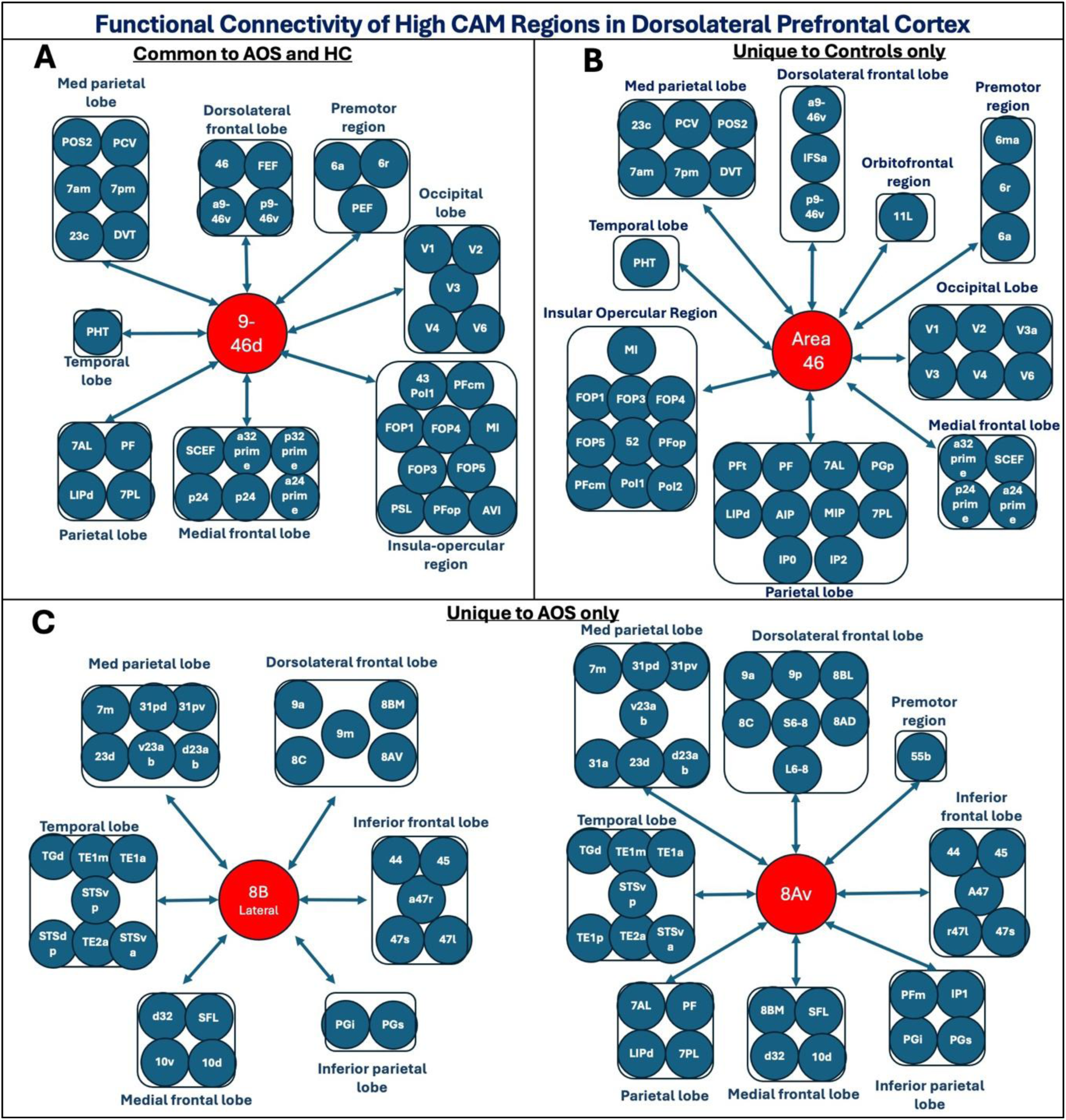
Functional connectivity of high CAM regions in the DLPFC Glasser section^1^. A. Functional Connectivity of Area 9-46d–one of the top 10% regions based on CAM weight for both AOS and controls. B. Another high CAM region (Area 46) in the DLFPC was unique to controls only. C. Area 8B Lateral and Area 8Av were unique to patients only. Since the classification of AOS was influenced by the functional connectivity of areas 9-46d, 8B lateral, and 8Av, compared to controls which was influenced by the functional connectivity of areas 9-46d and 46, dysfunction in more extensive networks of Glasser parcels in DLPFC section might have contributed to classification.

Other highly ranked class-activating nodes in both adult and AOS and respective controls were found in the visual and motor regions **(Supplemental Table 1 & 2).** This is consistent with recent meta-analysis that concluded that resting-state fMRI alterations are widespread but primarily include the visual, motor, and cerebellar systems, default-mode network, and striatum in first-episode psychosis^44^. Several default mode network nodes, identified on the Glasser atlas^45^, showed high CAM, especially for controls. These are the bilateral PGi, PFm, and POS2; these were in the top 10% of CAM nodes in both adolescent HC and adult HC. Notably, there were fewer default mode network-specific nodes to the patient class, besides the right PFm, which was in the top 10% of CAM nodes in all models and group. Right area 46 of the DLPFC, considered a part of the salience network^46^, was found to be in the top 10% of CAM nodes in AOS and adult schizophrenia.

High similarity of edgewise saliency by IG with CAM regions when IG is averaged by nodes (**Figure 5**) suggests that these features converge. Statistically similar nodal CAM weights and the regional sum of IG in all groups across both AOS and adult schizophrenia GCN models suggest that both saliency methods reveal comparably highly important regions and edges in the network for classification. Indeed, some key differences emerge when the IG saliency is not collapsed per node. First, for the resting state fMRI classification, connectivity in striatum was highly salient including left-to-right caudate in adolescent HC, the left-pallidum-to-left-putamen emerge in AOS (**Table 3**), and default mode network connectivity in AOS, AOS-controls, adult schizophrenia, and schizophrenia-controls, including area 10 and its subdivisions, and PGi/PGs. Second, the edgewise saliency values were normalized between zero and one on a whole-graph basis, whereas the CAM values are unbounded. Thus, a key point is the relative similarity of patterns of IG and CAM, as in **Figure 5**, not the exact value of the saliency magnitude, because many salient edges are required to constitute a feature used to inform classification.

For the working memory task, where the CAM and IG saliency are reported together (**Supplemental Table 4),** the cerebellum was important for working memory classification according to CAM, consistent with the “cerebellar cognitive theory” and implications for schizophrenia^40^. Previous evidence shows impaired working memory in schizophrenia^47^. We found significantly slower processing times for AOS by ≈100ms associated with the 1-back and by ≈200ms for the 2-back task with no groupwise differences in the accuracy of performance. We had reported similar findings in our prior study^48^. Accuracy of GCN classification was relatively low for even the best models for the working memory input set, i.e. 76.8%. Unlike the resting state analysis, saliency analysis could not be examined in the UK Biobank sample since working memory fMRI data was not collected, yet the results in **Supplemental Table 4** are consistent with previous neuroimaging literature of working memory in AOS, specifically, the association of Brodmann area 46 with working memory^49^. Aside from the sensory and motor areas, area 46 appears in the top 10 control and patient CAM regions. The more detailed perspective provided by IG indicates that the connectivity of Brodmann area 10 is highly influential for the classification; this region has been attributed in abstract cognition (decision making and problem solving) and working memory^1^, although we also found this edge to be salient for resting state classification (**Supplemental Table 4**).

There are limitations to the GCN training using dFCs. While non-stationarity, has been shown in both task-positive and task-free fMRI paradigms^50^ all acquisitions may not have detectable change-points. The ability to train a sufficiently accurate GCN as a prerequisite to saliency analysis can depend on sample size. Although our sample sizes for AOS and adult schizophrenia groups were relatively low, they are within the normal range of prior imaging studies^51^. However, with the implementation of the CCID algorithm, the number of networks used to train our GCN models appeared sufficient with over 300 networks in our smallest sample and over 3000 in the transdiagnostic sample. The similarity of salient features between adult schizophrenia and AOS samples demonstrates the robustness of our findings in different samples and separately trained models arguing for sufficient sample power.

This study illustrates the ability of a GCN classifier to be potentially used for clinical purposes. Accuracy of clinician’s psychiatric diagnoses is lower than our GCN model performance with accuracies as low as 55% in psychiatric diagnoses^52^ and 33% in schizophrenia spectrum disorders^53^. Since the reported reliability varied across the studies, we considered a clinician’s reliability of 65% to be conservative in estimating post-test probabilities. This can be achieved with a half-hour MRI scan after a clinician’s interview that is affordable in many countries. The GCN models also provide a biological interpretation to the classification through CAM and IG by identifying both regions and edges that can aid in further characterization as neuromodulation sites for therapy development^54^, especially in identifying specific targets for stimulation^55^. The results from our feature detection analyses could be an option to identify optimal disorder-specific or subject-specific stimulation sites.

## Methods

AOS and corresponding healthy control sample was locally acquired on a 7 Tesla whole-body scanner. The other was obtained from a publicly available 3T MRI dataset from the UK Biobank.

### Data Acquisition of UK Biobank

The UK Biobank (www.ukbiobank.ac.uk), is a large-scale database containing lifestyle and health data on about half a million participants in the United Kingdom^56^, including MRI imaging data for some participants^57^. The UK Biobank is a general population study and primarily includes participants with no ICD-10 disorder but also includes individuals with ICD-10 disorders including schizophrenia, bipolar disorder, and depression. UK Biobank data (accessed in March 2022) comprised of about 40,000 subjects with T_1_-weighted and rs-fMRI. Out of all subjects with required imaging modalities, we chose subjects meeting ICD-10 diagnostic criteria for schizophrenia (data field 130874), schizoaffective disorder (130884), manic episodes (130890), bipolar affective disorder (130892), and/or recurrent depressive disorder (130896). We grouped persons with these disorders together as the transdiagnostic group (n=127). The control group (n=127) comprised of individuals who were age-and-sex-matched to the transdiagnostic subjects. Image acquisition parameters have been previously documented^57,58^. T_1_w scans were acquired using 3D MPRAGE with voxel size=1 mm^3^. The rs-fMRI scans were acquired with voxel size=2.5mm^3^ and a TR=735ms.

### UK Biobank Preprocessing

The UK Biobank data were downloaded and processed using the Pittsburgh Supercomputing Center (PSC) Bridges-2 system^59^. The UK Biobank image files titled “T1_unbiased_brain.nii.gz”^58^ were processed with FreeSurfer without the high-resolution option. Resting state fMRI images were preprocessed by the UK Biobank^58^ for head motion (MCFLIRT)^60^, grand-mean intensity normalization of each 4D dataset, high-pass temporal filtering, echo planar imaging (EPI) unwarping and gradient distortion correction, and independent component analysis (ICA)-based denoising^61,62,63^.

### Data Acquisition of AOS

Participants were recruited from the Pittsburgh region between 2019 and 2023. Informed consent was obtained from all participants after describing the study, including the risks and benefits. The University of Pittsburgh Institutional Review Board approved the study. Subjects were administered the Structured Clinical Interview for DSM-IV (SCID-IV) and selected items on the Kiddie-Schedules for Assessment of Depression and Schizophrenia (K-SADS). Consensus diagnosis by medical investigators was made after reviewing all available clinical data including the charts. AOS is defined as onset psychotic symptom after puberty, but before completing 18 years of age. Healthy controls were also recruited from the same geographic region.

### AOS Imaging Methods

The AOS MRI data included structural MRI, and both a working memory fMRI and rs-fMRI. MR images were collected on a Siemens Magnetom whole-body 7T scanner using a custom 64-channel Tic-Tac-Toe head coil. T1-weighted MP2RAGE scans were acquired in the axial plane using 348 slices with 0.55 mm thickness, a TE=2.54ms, TR=6 seconds, and an in-plane resolution=390×390. The rs-fMRI data was acquired during an 8-minute scan when subjects were observing a fixation cross and task-fMRI acquired during a letter N-back working memory task.

The N-back task made use of a response glove to gather subject responses. The task was presented using the E-Prime 2 software. Six task blocks were presented interleaved by fixation blocks. The scan began with a task block lasting 32 TR (80 seconds), followed by a fixation block of 8 TR (20 seconds). The stimuli were letters from the alphabet. The task blocks were either 0-back, where the participants are expected to respond only to the letter “x,” 1-back, where the target response is any letter shown twice in a row, and 2-back, where the target response is any letter shown two stimuli ago. The response window was 2.5 seconds (one TR length). Each block contained 6 target letters among 32 stimuli. Across all blocks there was a total of 36 possible correct, with positions and quality of stimulus randomized between subjects, as well as a random block order.

### AOS MRI Data Preprocessing

Freesurfer processing with `recon-all` included the high-resolution option recommended for images with less than 1mm isotropic resolution. All fMRI preprocessing steps were performed using fMRIPrep^64^, and included correction for rigid body-head motion, slice timing, bias, gradient distortion, and finally brain extraction all under the guidelines of the Human Connectome Project principal of minimal preprocessing^65^.

### Atlas-based fMRI Timeseries Extraction

Anatomical image preprocessing consists of denoising, tissue labeling, and co-registration to fMRI images. FreeSurfer (version 7.2.0)^66^ was used for preprocessing and segmentation with the `recon-all` function, as described in the sections above. Afterwards, each individual brain was parcellated according to the Human Connectome Project Multi-Modal Parcellation (HCP-MMP1) atlas^35^, using a published method (NeuroLab). Briefly, this method uses a Freesurfer average version of the HCP MMP1 parcellation to map to the individual result of `recon-all` and then create a volumetric representation of the standard 180-region-per-hemisphere Glasser parcellation in native space. Two of the Glasser atlas regions are the left and right hippocampus. These are included among the Freesurfer-derived subcortical structures, so we consider there to be 358 *cortical* parcels, plus the 27 subcortical parcels (which included bilateral hippocampi). However, the left and right substantia nigra were too small to be reliably parcellated using the automated segmentation. These were removed to yield 25 subcortical structures, resulting in a total of 383 regions.

After fMRI and anatomical preprocessing, registration of anatomical volumes to fMRI space was performed and then the transform applied to the atlas using nearest neighbor interpolation to provide fMRI-space atlases using the FSL linear alignment tool^60^. For each brain region (parcel) in the registered atlas image, the voxel-wide average signal activation value of the preprocessed fMRI was calculated for each parcel at each timepoint. This resulted in a multivariate time series of size equal to the number of nodes in the brain atlas by the number of fMRI timepoints. For each acquisition, each nodal timeseries was then *z*-scored per-node over time^67^ to normalize the values of fMRI signal.

### Change-point Detection

CCID uses the isolate-detect principle^31^ to estimate the number and temporal positions of change-points in a multivariate timeseries^32^. The algorithm converts the multivariate timeseries into local wavelet periodograms and cross-periodograms, and aggregates across the timeseries to simplify the problem in high dimensional data.

CCID requires the experimenter to define parameters which will influence the estimate. Four primary configurations of CCID were compared^32^, all of which outperformed existing change-point detection algorithms across several simulation datasets. These four configurations represent top-level choices of change-point detection methods. The first choice pertains to change-point detection (pruning) and can be either “threshold” or “information criterion”, abbreviated TH and IC. For detection, we used the IC method because it is considered an advancement over the TH method. The second choice is of the metric used for multivariate timeseries aggregation, and is either the Euclidean approach, *L_2_*, or the “infinity” approach, *L_∞_*. We used the Euclidean approach since we found it to be faster than and equivalent to the infinity approach; additionally, Anastasiou et al noted that “the *L_2_* threshold works better for small changes across many [time]series”^32^ which is expected in fMRI data, where signal-to-noise is low and timeseries are high dimensional.

Small adjustments in the parameter choices of the CCID function were made for each dataset. The wavelet scale was found to be an important factor for change-point detection. The wavelet scale parameter in the CCID function is valued for negative integers, where small negative values correspond to smaller scales (higher frequencies), while large negative values correspond to larger scales (lower frequencies). By evaluating the number of change-points detected at a range of different wavelet scales, we have determined the wavelet scale parameter uniquely for each dataset. For the UK Biobank sample, a wavelet scale of –4 was used, whereas for the AOS dataset, a scale of –3 was used. In both datasets at these scale values, nearly all subjects had more than zero change-points detected. This parameter is associated with the expected frequency “packets” in the fMRI timeseries, and thus can be dependent on stimulus design and, more importantly, sampling rate of the fMRI. A minimum distance between change-points (δ value) was specified for the UK Biobank of 10, whereas in the AOS dataset no minimum distance was set but dynamic windows of less than 10 were not considered. Because the UK Biobank dataset was denoised using ICA, all windows were considered to be without motion; whereas with the AOS dataset, motion windows were identified where framewise displacement was over 2.5mm. When framewise displacement was more than 2.5mm, those dFC windows were discarded.

### Identification of dFCs from Change-points

Pearson correlation was used to calculate connectivity between region-pairs of preprocessed fMRI signal for each window between neighboring change-points, detected using the CCID algorithm. For example, in **Figure 1C**, red lines in the middle row represent where a change-point was detected and the networks were calculated using the signals between the two change-points. The first dFC was defined from the start of the task to the first change-point and the last dFC was defined using the signal from the last change-point to the end of the scan.

Since FC matrices are complete graphs, edge selection methods need to be employed to create sparse graphs. Many edge selection methods exist to eliminate edges with low statistical probability, with little consensus about the “correct” thresholding method^68^. We have chosen to use statistical thresholding because it is commonly used^69^ and simply relies on the *p*-value of the fMRI correlation at each edge between two brain regions (or nodes), independently. Binary graphs were created where any edge with a p-value greater than 0.05 was set to zero and edges with nominally significant p-values were set to 1, regardless of the magnitude of the correlation.

### GCN Architecture and Training

We customized^16^ a published GCN pipeline^15^ to examine dFC classification (**Figure 3**). Modifications were made to the type of edge selection method and input network size, and therefore how the data was loaded into Pytorch. Each input to our model was a 383×383 unweighted dFC matrix where the nodes are regions of the brain and node features are the connectivity between that region and all other regions. This connectivity/adjacency matrix was input into the GCN model using a tensor which consisted of the adjacency matrix, and the edge locations. In short, three graph convolutional layers (number of channels: [64, 64, 128]) were used in our model followed by a global average pooling layer and a fully connected layer. Each convolutional layer used Chebyshev convolution with *k* order=12. The Rectified Linear Unit (ReLU) activation function was used to add hidden layers and introduce non-linearity into the model. The softmax function encoded the output value into a predictive probability for each class from the output layer.

To mitigate overfitting, we implemented several strategies. First, we employed weight decay (5e-4) in the Adam optimizer, which acts as *L_2_* regularization to penalize large weights. Second, we utilized a stratified *k*-fold cross-validation approach *(k*=5 or *k*=10 for transdiagnostic sample only) to ensure consistent class distribution across all folds with the test-validation set consisting of 80% of the entire sample and the test set was the remaining 20%. Within each fold, the training-validation set was further stratified (89% training/11% validation) to maintain class balance. Most importantly, we implemented an early stopping mechanism where model performance was continuously monitored on the validation set. The best model for each fold was saved based on the lowest validation loss rather than training metrics, preventing the model from overfitting to the training data. This approach resulted in models that generalize better to unseen data, as evidenced by the balanced test metrics across folds. For each fold, up to 50 epochs of the training dataset were run through the learning process; the final model selected was the one with minimum validation loss. The performance was evaluated by calculating training loss, validation loss, balanced accuracy, sensitivity, and specificity on both validation and test sets.

### Salient Feature Detection

To investigate the salient features, or the features of the connectomes which were most important for classification, we implemented two methods, namely Class Activation Mapping (CAM) and Integrated Gradients (IG) (**Figure 3**). CAM detects salient regions (nodes), and integrated gradients detect salient edges within the networks. When reporting these results, we used the best performing model in terms of classification metrics, from the resting-state for the AOS and the UK Biobank adult schizophrenia models because these were our most comparable groups as well as the samples with the highest accuracy. We investigated their salient features (regions and edges), separately and also compared them to detect similar salient features across both groups. Working memory salient features for the AOS dataset are also reported.

### Class Activation Mapping

After training, we used CAM to detect salient regions^17^. Using the GCN model with the highest accuracy, we extracted the final dense layer, or the output from the final convolutional layer, from all subjects. To estimate the activation value of each node, we multiplied each subject’s final layer (in our case 383×128 matrix) by the optimized weight vector for their respective true class (128×1 vector). This yielded a 383×1 CAM vector for each dFC in each class. Here, we were interested in analyzing the two groups’ activation values separately, so we calculated the average CAM vector across subjects in each case-control group. The top ten percent of regions (38 regions) ranked in terms of high to low activation weights are reported for each group for conciseness, but the rank of all regions can be found in the supplemental spreadsheet.

### Integrated Gradients

We used IG to identify the most salient edges within the networks^70^ by using the GCN model with the highest accuracy to analyze the contribution of each edge to the classification outcome. For each subject, the edge importance scores were computed by integrating the gradients of the model’s output with respect to the input edge weights along a path from a baseline (zero-edge connectivity) to the input graph using the Integrated Gradients class from the Captum library of the PyTorch. This yielded a salience matrix for each dFC, where each cell represented the importance of a specific edge. This matrix had a size of 383×383 (representing all connections), where each element *(i,j*) corresponded to the salience score of the edge between regions *i* and *j*.

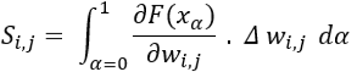

Here, *S_i,j_* represents the importance of the edge between nodes *i* and *j*. The model’s prediction function, *F(x_α_)*, was applied to the interpolated graph *x_α_*, which transitions from a baseline graph (with no edge weights) to the input graph. The gradient *∂F (x_α_)/∂w_i,j_* quantifies the sensitivity of the model’s output to changes in the edge weight *w_i,j_*, while *Δ w_i,j_* captures the difference between the actual edge weight and the baseline (of zero). The parameter *α* interpolates between the baseline (where *α*=0) and the input graph (where *α*=1). The integral over *α* aggregates the gradients along this path, providing a comprehensive measure of each edge’s contribution to the model’s prediction. This method ensures that the calculated salience scores accurately reflect the influence of specific edges within the network.

The salience scores were normalized so that the maximum value was scaled to 1, making it easier to compare edge importance across subjects and identify consistently influential edges. To identify group-level patterns, the average salience matrix for each group was computed. The top-ranked edges, based on their importance scores, were identified, and reported for each group. The salience matrices for different groups were compared to identify both shared and unique salient edges.

### Statistics

For all five GCN input sets, AOS-working memory, AOS-resting, UKB-Adult schizophrenia, UKB-Bipolar, UKB-Major Depression, and a sixth amalgamated UKB-Transdiagnostic, the number of change-points per subject was calculated and the population distributions of the number of change-points over all subjects within a patient group were compared to the population distribution of number of change-points per subject among corresponding controls using a two-sample Kolmogorov-Smirnov test (KS-test). Similarly, patient-control differences in the population distributions of dFC window lengths between two sequential change-points were also measured using a two-sample KS-test. For the AOS study only, which is a block-design working memory fMRI task, change-points can be considered “expected” if they happen near a change from a task block to a fixation block (or from fixation to task block), or they can be considered “unexpected” if they happen mid-task or mid-fixation. The proportion of expected versus unexpected change-points between the patient and control groups was compared using a chi-squared test of proportions, where change-points were considered expected if they happened at the first TR of a new block, t_0_, or no more than 2 TR before or after t_0_. GCN performance was compared using both best model performance across all folds and average model performance.

For the three input sets containing schizophrenia comparisons, which were the AOS working memory, AOS resting state, and UK Biobank adult schizophrenia resting state, the best models were selected for saliency analysis. CAM was calculated for all nodes and the top 10% of class activation nodes were identified in each input set, for each class. The top 10 salient network edges were also listed (much less than 1% of all edges). For the input set containing the N-back working memory task, groupwise differences in task performance in terms of total correct and response times in milliseconds for 0-back, 1-back, 2-back, and whole-task were assessed using two-sample T-tests.

Further comparisons were made between the two input sets that were most similar, namely the AOS resting state and the UKB adult schizophrenia. The top 10% of nodes (38 nodes) were projected onto 3D renderings of the brain to demonstrate spatial overlap of a broad swath of class activating nodes between patients and controls as well as between AOS and adult schizophrenia (between input sets, i.e. comparing different models). The linear relationship between the intra-class CAM and nodal average IG was assessed using Pearson’s correlation (*r*) and adjusted *R*^2^. To compare the saliency results from different GCN models trained on independent samples, the same statistics were used to assess the linear relationship of CAM values across nodes between AOS and adult schizophrenia, as well as nodal average IG values across nodes between AOS and adult schizophrenia; equivalent healthy control comparisons were also performed between the adolescent controls and the adult controls for both CAM and IG.

## Data Availability

UK Biobank available upon application to the Biobank. Locally collected data available with the NDA

## Acknowledgements

We want to thank the Pittsburgh Supercomputing Center for providing the computational resources through ACCESS Discover grant BIO200047. Additionally, we would like to thank UK Biobank for providing the data for this study.

---

**Supplemental Figure 1:**
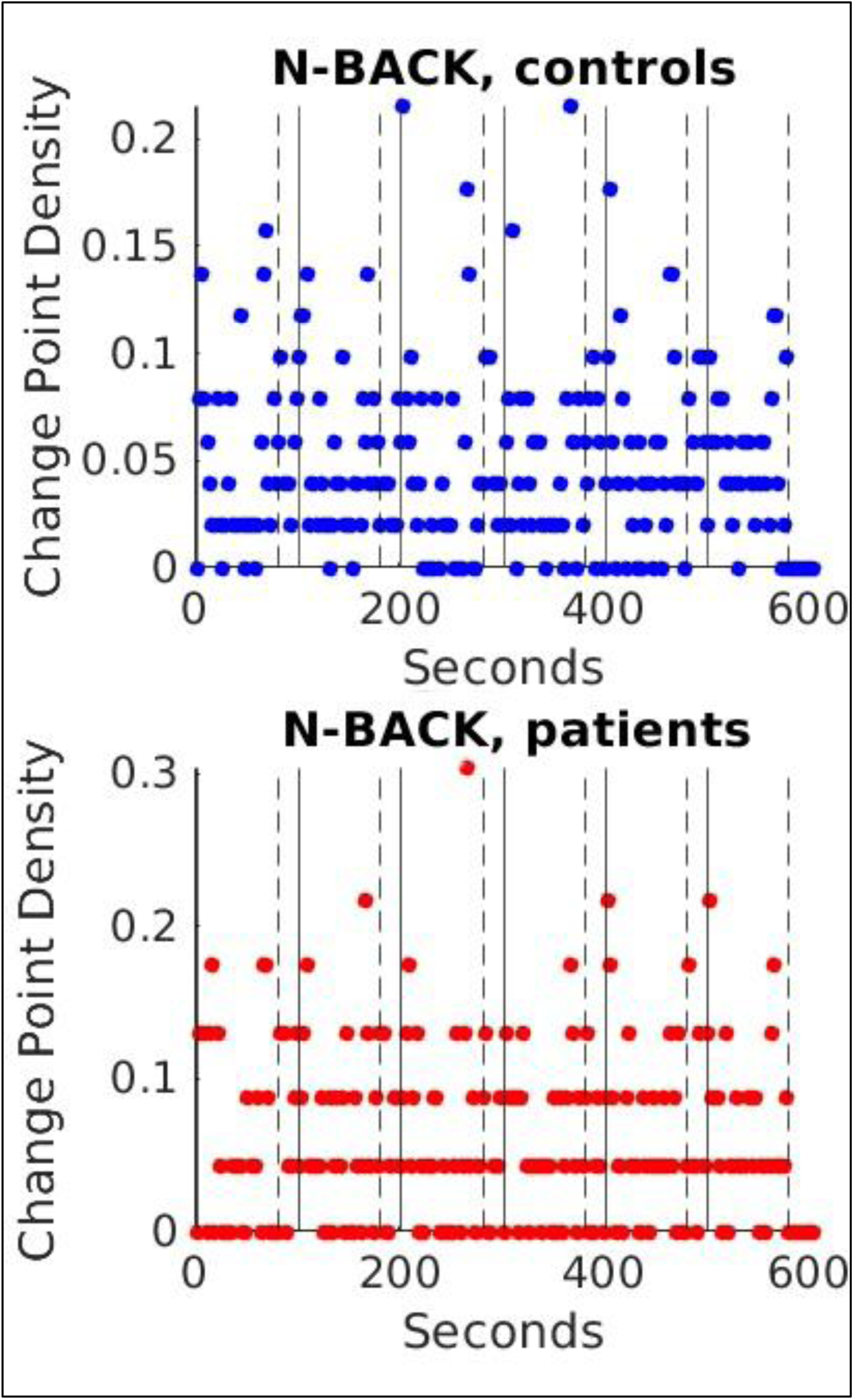
Density of changepoints detected by CCID method plotted against time on N-back task fMRI data among AOS and HC. The top plot represents controls (blue scatter points) and the bottom plot represents patients (red). Scatter plots are shown for all subjects. In both plots, the x-axis is time (in seconds) since the start of the fMRI acquisition and the y-axis represents the “change point density,” or population frequency of changepoints at that time. Dashed vertical lines indicate the onset of the inter-trial fixation block, and solid black lines the onset of the task block. The acquisition begins with an N-back task block and ends with fixation block.

**Supplemental Table 1:** AOS vs control model regions with CAM weights in the top 10%. Regions with a blue fill are common to both groups, green fill is unique to the patient group, and yellow fill is unique to control group.

| Region Name | Glasser Section |
| --- | --- |
| Regions common to both patient and control groups |  |
| Left-Cerebellum-Cortex | subcortical |
| Right-Cerebellum-Cortex | subcortical |
| Left Primary Visual Cortex | Primary Visual Cortex |
| Left Second Visual Area | Early Visual Cortex |
| Left Third Visual Area | Early Visual Cortex |
| Left Primary Motor Cortex | Somatosensory and Motor Cortex |
| Left Primary Sensory Cortex | Somatosensory and Motor Cortex |
| Left Medial Area 7A | Superior Parietal Cortex |
| Left Area 1 | Somatosensory and Motor Cortex |
| Right Primary Visual Cortex | Primary Visual Cortex |
| Right Second Visual Area | Early Visual Cortex |
| Right Third Visual Area | Early Visual Cortex |
| Right Primary Motor Cortex | Somatosensory and Motor Cortex |
| Right Primary Sensory Cortex | Somatosensory and Motor Cortex |
| Right Area V3A | Dorsal Stream Visual Cortex |
| Right Area 7m | Posterior Cingulate Cortex |
| Right Medial Area 7A | Superior Parietal Cortex |
| Right Area 9-46d | Dorsolateral Prefrontal Cortex |
| Right Auditory 5 Complex | Auditory Association Cortex |
| Right Area PHT | Lateral Temporal Cortex |
| Right Area PFm Complex | Inferior Parietal Cortex |
| Right Area PGI | Inferior Parietal Cortex |
| Regions unique to the patient group |  |
| Left Area V3A | Dorsal Stream Visual Cortex |
| Left Parieto-Occipital Sulcus Area 2 | Posterior Cingulate Cortex |
| Left Area 8Av | Dorsolateral Prefrontal Cortex |
| Left Area 6 anterior | Premotor Cortex |
| Left Area PGp | Inferior Parietal Cortex |
| Left Area PFm Complex | Inferior Parietal Cortex |
| Right Fourth Visual Area | Early Visual Cortex |
| Right Parieto-Occipital Sulcus Area 2 | Posterior Cingulate Cortex |
| Right Superior Temporal Visual Area | Temporo-Parieto-Occipital Junction |
| Right Area 8B Lateral | Dorsolateral Prefrontal Cortex |
| Right Area 10v | Anterior Cingulate and Medial Prefrontal Cortex |
| Right Area TE1 posterior | Lateral Temporal Cortex |
| Right Area TemporoParietoOccipital Junction 1 | Temporo-Parieto-Occipital Junction |
| Right Dorsal Transitional Visual Area | Posterior Cingulate Cortex |
| Right Area PGp | Inferior Parietal Cortex |
| Right Area IntraParietal 2 | Inferior Parietal Cortex |
| Regions unique to the control group |  |
| Left Lateral Area 7A | Superior Parietal Cortex |
| Left Dorsal area 6 | Premotor Cortex |
| Left Area 6mp | Paracentral Lobular and Mid Cingulate Cortex |
| Left Area 9-46d | Dorsolateral Prefrontal Cortex |
| Left Auditory 5 Complex | Auditory Association Cortex |
| Left Area STSd anterior | Auditory Association Cortex |
| Right Frontal Eye Fields | Premotor Cortex |
| Right Area 6m anterior | Paracentral Lobular and Mid Cingulate Cortex |
| Right Area 7PC | Superior Parietal Cortex |
| Right Area 1 | Somatosensory and Motor Cortex |
| Right Area 2 | Somatosensory and Motor Cortex |
| Right Dorsal area 6 | Premotor Cortex |
| Right Area 6mp | Paracentral Lobular and Mid Cingulate Cortex |
| Right Area 46 | Dorsolateral Prefrontal Cortex |
| Right Area PFt | Inferior Parietal Cortex |
| Right Auditory 4 Complex | Auditory Association Cortex |

**Supplemental Table 2:** SZ vs control model regions with CAM weights in the top 10%. Regions with blue fill are common to both groups, green fill is unique to the patient group, and yellow fill is unique to control group.

| Region Name | Glasser Section |
| --- | --- |
| Regions common to both patient and control groups |  |
| Left Cerebellum Cortex | Subcortical |
| Right Cerebellum Cortex | Subcortical |
| Left Primary Visual Cortex | Primary Visual Cortex |
| Left Second Visual Area | Early Visual Cortex |
| Left Third Visual Area | Early Visual Cortex |
| Left Fourth Visual Area | Early Visual Cortex |
| Left Primary Motor Cortex | Somatosensory and Motor Cortex |
| Left Parieto-Occipital Sulcus Area 2 | Posterior Cingulate Cortex |
| Left Medial Area 7A | Superior Parietal Cortex |
| Left Lateral Area 7P | Superior Parietal Cortex |
| Left Area PHT | Lateral Temporal Cortex |
| Left Area IntraParietal 1 | Inferior Parietal Cortex |
| Left Area PF Complex | Inferior Parietal Cortex |
| Left Area PFm Complex | Inferior Parietal Cortex |
| Left Auditory 4 Complex | Auditory Association Cortex |
| Right Primary Visual Cortex | Primary Visual Cortex |
| Right Second Visual Area | Early Visual Cortex |
| Right Third Visual Area | Early Visual Cortex |
| Right Parieto-Occipital Sulcus Area 2 | Posterior Cingulate Cortex |
| Right Medial Area 7A | Superior Parietal Cortex |
| Right Area 46 | Dorsolateral Prefrontal Cortex |
| Right Area 9-46d | Dorsolateral Prefrontal Cortex |
| Right Anterior IntraParietal Area | Superior Parietal Cortex |
| Right Area PHT | Lateral Temporal Cortex |
| Right Area PGp | Inferior Parietal Cortex |
| Right Area IntraParietal 1 | Inferior Parietal Cortex |
| Right Area PF Complex | Inferior Parietal Cortex |
| Right Area PFm Complex | Inferior Parietal Cortex |
| Right Auditory 4 Complex | Auditory Association Cortex |
| Regions unique to the patient group |  |
| Left Area V3A | Dorsal Stream Visual Cortex |
| Left Area 7m | Posterior Cingulate Cortex |
| Left Area TE1 posterior | Lateral Temporal Cortex |
| Left Area PGI | Inferior Parietal Cortex |
| Left Area PGs | Inferior Parietal Cortex |
| Right Fourth Visual Area | Early Visual Cortex |
| Right Area V3A | Dorsal Stream Visual Cortex |
| Right Area PGI | Inferior Parietal Cortex |
| Right Area PGs | Inferior Parietal Cortex |
| Regions unique to the control group |  |
| Left IntraParietal Sulcus Area 1 | Dorsal Stream Visual Cortex |
| Left Medial Area 7p | Superior Parietal Cortex |
| Left Area anterior 9-46v | Dorsolateral Prefrontal Cortex |
| Left Area PGp | Inferior Parietal Cortex |
| Area IntraParietal 2 | Inferior Parietal Cortex |
| Right Medial IntraParietal Area | Superior Parietal Cortex |
| Right Area 2 | Somatosensory and Motor Cortex |
| Right Rostral Area 6 | Premotor Cortex |

**Supplemental Table 3:** Statistical differences between AOS patient and control groups for N-back accuracy (A) and processing time (B). Average scores show mean and standard deviation with the overall score being out of 36 and 0-back,1-back, and 2-back out of 12.

| A. Statistical Differences in N-back Accuracy |  |  |  |  |
| --- | --- | --- | --- | --- |
|  | AOS score | Control score | t | p-value |
| Overall | 30.0 ± 6.4 | 32.4 ± 5.5 | 1.62 | p>0.05 |
| 0-back | 10.9 ± 2.8 | 11.2 ± 2.1 | 0.53 | p>0.05 |
| 1-back | 10.4 ± 2.4 | 11.2 ± 1.9 | 1.56 | p>0.05 |
| 2-back | 8.7 ± 2.4 | 9.9 ± 2.7 | 1.85 | p>0.05 |
| B. Statistical Differences in N-back Processing Time |  |  |  |  |
|  | AOS | Control | t | p-value |
| Overall | 710.0 ± 220.9 ms | 601.3 ± 90.6 ms | 1.62 | p<0.01 |
| 0-back | 534.2 ± 87.4 ms | 527.0 ± 73.1 ms | 0.35 | p>0.05 |
| 1-back | 727.5 ± 298.5 ms | 614.8 ± 108.0 ms | 1.86 | p<0.05 |
| 2-back | 889.5 ± 350.1 ms | 666.2 ± 169.0 ms | t=3.59 | p<0.001 |

**Supplemental Table 4:** Top 10% salient CAM regions by class, and top 10 salient IG connections by class for working memory derived dFCs. Red font indicates a similar region or connection between the control and patient groups.

| Top 10 Working Memory Salient Nodes – Working Memory by CAM |  |  |  |  |
| --- | --- | --- | --- | --- |
|  | Controls |  | Adolescent- Onset Schizophrenia |  |
| Rank | Region Name | Glasser Section | Region Name | Glasser Section |
| 1 | L Cerebellum Cortex | Subcortical | R Cerebellum Cortex | Subcortical |
| 2 | R Primary Motor Cortex | Somatosensory and Motor Cortex | L Cerebellum Cortex | Subcortical |
| 3 | R Cerebellum Cortex | subcortical | L Primary Motor Cortex | Somatosensory and Motor Cortex |
| 4 | L Area 9-46d | Dorsolateral Prefrontal Cortex | R Primary Sensory Cortex | Somatosensory and Motor Cortex |
| 5 | R Second Visual Area | Early Visual Cortex | R Area 7PC | Superior Parietal Cortex |
| 6 | R Area 46 | Dorsolateral Prefrontal Cortex | R Area 46 | Dorsolateral Prefrontal Cortex |
| 7 | L Primary Motor Cortex | Somatosensory and Motor Cortex | L Primary Visual Cortex | Primary Visual Cortex |
| 8 | R Anterior 24 prime | Anterior Cingulate and Medial Prefrontal Cortex | L Area 9-46d | Dorsolateral Prefrontal Cortex |
| 9 | R Area 9-46d | Dorsolateral Prefrontal Cortex | L Medial Area 7A | Superior Parietal Cortex |
| 10 | L Primary Sensory Cortex | Somatosensory and Motor Cortex | R Primary Visual Cortex | Primary Visual Cortex |
| 11 | R Frontal Eye Fields | Premotor Cortex | R Area 2 | Somatosensory and Motor Cortex |
| 12 | R Area 2 | Somatosensory and Motor Cortex | L Area dorsal 32 | Anterior Cingulate and Medial Prefrontal Cortex |
| 13 | L Parieto-Occipital Sulcus Area 2 | Posterior Cingulate Cortex | L Area 1 | Somatosensory and Motor Cortex |
| 14 | R Area 1 | Somatosensory and Motor Cortex | L Area posterior 24 | Anterior Cingulate and Medial Prefrontal Cortex |
| 15 | L Second Visual Area | Early Visual Cortex | R Primary Motor Cortex | Somatosensory and Motor Cortex |
| 16 | R PreCuneus Visual Area | Posterior Cingulate Cortex | L Second Visual Area | Early Visual Cortex |
| 17 | R Area 55b | Premotor Cortex | L Area 2 | Somatosensory and Motor Cortex |
| 18 | R Medial Area 7A | Superior Parietal Cortex | L Primary Sensory Cortex | Somatosensory and Motor Cortex |
| 19 | R Superior Temporal Visual Area | Temporo-Parieto-Occipital Junction | L Third Visual Area | Early Visual Cortex |
| 20 | R Primary Sensory Cortex | Somatosensory and Motor Cortex | R Second Visual Area | Early Visual Cortex |
| 21 | R Rostral Area 6 | Premotor Cortex | L Area 46 | Dorsolateral Prefrontal Cortex |
| 22 | R Parieto-Occipital Sulcus Area 2 | Posterior Cingulate Cortex | R Area 9-46d | Dorsolateral Prefrontal Cortex |
| 23 | L Area 2 | Somatosensory and Motor Cortex | R Auditory 5 Complex | Auditory Association Cortex |
| 24 | L Primary Visual Cortex | Primary Visual Cortex | R Area 1 | Somatosensory and Motor Cortex |
| 25 | R Area 8Av | Dorsolateral Prefrontal Cortex | L Area 9 Middle | Anterior Cingulate and Medial Prefrontal Cortex |
| 26 | R Area 6mp | Paracentral Lobular and Mid Cingulate Cortex | L Area PFm Complex | Inferior Parietal Cortex |
| 27 | L Area Posterior 24 prime | Anterior Cingulate and Medial Prefrontal Cortex | L Fourth Visual Area | Early Visual Cortex |
| 28 | L Area 1 | Somatosensory and Motor Cortex | L Area 8BM | Anterior Cingulate and Medial Prefrontal Cortex |
| 29 | L Area PF Complex | Inferior Parietal Cortex | R Anterior IntraParietal Area | Superior Parietal Cortex |
| 30 | R Area 8BM | Anterior Cingulate and Medial Prefrontal Cortex | L Area 45 | Inferior Frontal Cortex |
| 31 | R Supplementary and Cingulate Eye Field | Paracentral Lobular and Mid Cingulate Cortex | R Superior Frontal Language Area | Dorsolateral Prefrontal Cortex |
| 32 | R Ventral Area 6 | Premotor Cortex | L Area 6m anterior | Paracentral Lobular and Mid Cingulate Cortex |
| 33 | L Area 46 | Dorsolateral Prefrontal Cortex | R Area 8Av | Dorsolateral Prefrontal Cortex |
| 34 | R Primary Visual Cortex | Primary Visual Cortex | R Area 8C | Dorsolateral Prefrontal Cortex |
| 35 | R Area Posterior 24 prime | Anterior Cingulate and Medial Prefrontal Cortex | L Area 7PC | Superior Parietal Cortex |
| 36 | L Area anterior 9-46v | Dorsolateral Prefrontal Cortex | L Dorsal Area 24d | Paracentral Lobular and Mid Cingulate Cortex |
| 37 | R Area TemporoParietoOccipital Junction 1 | Temporo-Parieto-Occipital Junction | R Third Visual Area | Early Visual Cortex |
| 38 | R Area 23c | Paracentral Lobular and Mid Cingulate Cortex | L Area IFSa | Inferior Frontal Cortex |

Top 10 Salient Working Memory Connections of Adolescent-Onset Schizophrenia Model
|  | Controls |  | Schizophrenia |  |
| --- | --- | --- | --- | --- |
| Rank | Node 1 | Node 2 | Node 1 | Node 2 |
| 1 | R Area V4t | R Area Lateral Occipital 2 | R Area 10d | R Area 9 anterior |
| 2 | R Anterior IntraParietal Area | R Area IntraParietal 2 | R Area 9 anterior | R Area 10d |
| 3 | L Area 9 anterior | L Area 9 Posterior | R Area 10d | L Area 10d |
| 4 | L Orbital Frontal Complex | L Area 10v | L Area 10d | R Area 10d |
| 5 | R Area 10d | L Area 10d | L Lateral Belt Complex | L ParaBelt Complex |
| 6 | L Area ventral 23 a+b | R Area ventral 23 a+b | L Area Lateral Occipital 2 | L Area V4t |
| 7 | L Area 8B Lateral | R Area 8B Lateral | R Orbital Frontal Complex | L Orbital Frontal Complex |
| 8 | R Area 10v | R Area 10r | L Area V4t | L Area Lateral Occipital 2 |
| 9 | L Polar 10p | L Area 10v | R RetroSplenial Complex | L RetroSplenial Complex |
| 10 | L Orbital Frontal Complex | R Orbital Frontal Complex | R Area 8B Lateral | L Area 8B Lateral |

